# Oral antiviral clevudine compared with placebo in Korean COVID-19 patients with moderate severity

**DOI:** 10.1101/2021.12.09.21267566

**Authors:** Joon-Young Song, Yeon-Sook Kim, Joong-Sik Eom, Jin-Yong Kim, Jin-Soo Lee, Jacob Lee, Won-Suk Choi, Jung-Yeon Heo, Jang-Wook Sohn, Ki-Deok Lee, Donghui Cho, Ilyoung Cho, Woo-Joo Kim

**Affiliations:** Division of Infectious Diseases, Department of Internal Medicine, Korea University College of Medicine, Seoul, Korea; Division of Infectious Diseases, Department of Internal Medicine, Chungnam National University School of Medicine, Daejeon, Republic of Korea; Division of Infectious Diseases, Department of Internal Medicine, Gil Medical Center, Gachon University School of Medicine, Incheon, Republic of Korea; Division of Infectious Diseases, Department of Internal Medicine, Incheon Medical Center, Incheon, Republic of Korea; Division of Infectious Diseases, Department of Internal Medicine, Inha University School of Medicine, Incheon, Republic of Korea; Division of Infectious Disease, Department of Internal Medicine, Kangnam Sacred Heart Hospital, Hallym University College of Medicine, Seoul, Republic of Korea; Division of Infectious Diseases, Department of Internal Medicine, Korea University College of Medicine, Ansan Hospital, Ansan, Republic of Korea; Department of Infectious Diseases, Ajou University School of Medicine, Suwon, Korea; Division of Infectious Diseases, Department of Internal Medicine, Korea University College of Medicine, Seoul, Republic of Korea; Department of Infectious Diseases, Myongji Hospital, Hanyang university medical center, Goyang, Republic of Korea; Department of Surgery, Seoul Medical Center, Seoul, Republic of Korea; Bukwang Pharm. Co. Ltd., Seoul, Republic of Korea

## Abstract

**Background:** Clevudine, an antiviral drug for chronic hepatitis B virus infection, is expected to inhibit the replication of severe acute respiratory syndrome coronavirus 2 (SARS-CoV-2) virus. Therefore, we conducted a prospective, single-blind, proof of concept clinical study to examine the antiviral efficacy and safety of clevudine compared to placebo in Korean corona virus disease 19 (COVID-19) patients with moderate severity.

**Methods:** Adults with confirmed SARS-CoV-2 infection and symptom onset within 7 days were randomized 2:1 to 120 mg clevudine or placebo to receive one of treatments orally once-daily for 14 days. Antiviral efficacy outcomes were the proportion of patients with real-time reverse transcription polymerase chain reaction (RT-PCR) negative result for SARS-CoV-2 infection and cycle threshold (Ct) value changes from baseline. Clinical efficacy outcomes included proportion of patients who showed improvement in lung involvement by imaging tests, proportion of patients with normal body temperature, proportion of patients with normal oxygen saturation, and the changes in C-reactive protein (CRP) from baseline. Safety outcomes included changes in clinical laboratory tests, vital signs measurement, and physical examination from baseline, and incidence of adverse events.

**Results:** The proportion of patients with real-time RT-PCR negative test and Ct value changes showed no significant difference between clevudine group and placebo group. The changes in Ct value from baseline were significantly greater in clevudine group compared to placebo group in patients with hypertension, and patients who underwent randomization during the first 5 and 7 days after the onset of symptoms. All clinical efficacy outcomes had no significant difference between clevudine group and placebo group. Clevudine was well tolerated and there was no significant difference in safety profile between two treatment groups.

**Conclusions:** This is the first clinical study to compare the antiviral efficacy and safety of clevudine to placebo in Korean COVID-19 patients with moderate severity. The study has demonstrated a possible favorable outcome for the reduction of SARS-CoV-2 replication, with acceptable safety profile, when COVID-19 patients were treated with clevudine. Further large-scale clinical studies, preferably with various clinical endpoints and virus titer evaluation, are required to better understand the effectiveness of using clevudine in COVID-19 treatment. Considering recent trend in clinical development for antiviral drugs, we need to design a clinical study aiming for reducing clinical risk of COVID-19 in mild to moderate patients with at least one risk factor for serious illness.

## Introduction

A new species of coronavirus, severe acute respiratory syndrome coronavirus 2 (SARS-CoV-2), first identified at Wuhan city of China in December 2019, causes the respiratory infection designated as corona virus disease 19 (COVID-19) has led to a global pandemic as announced by the World Health Organization (WHO) on 11 March 2020.^1^ Clinical symptoms of COVID-19 range from asymptomatic case to respiratory illness (pneumonia) with fever, sneeze, cough, and shortness of breath.^2^ Several demographic factors and comorbidities (e.g., hypertension, diabetes mellitus, and liver disease) are known to be associated with worse outcomes in COVID-19.^3-5^ In particular, hypertension is demonstrated as one of the high-risk factors associated with aggravation of severity and higher mortality in patients with COVID-19.^6, 7^ It is well known that SARS-CoV-2 viral load peaks at the time of symptom onset, and gradually decreases soon after.^8^ Based on this knowledge, various clinical trials and research studies of COVID-19 treatments have been designed to elucidate the effect of treatment initiation timing from symptom onset in COVID-19 management.^9-11^

SARS-CoV-2 is a single-stranded positive-sense RNA virus whose cell entry is mediated by the binding of the virus spike protein to angiotensin-converting enzyme 2 (ACE2), an enzyme attached to the membrane of cells. Subsequent viral-host membrane fusion allows the release of the viral RNA into the host cells. The 3CL protease (3CLpro), known as the main protease of the SARS-CoV-2, plays a major role to produce non-structural proteins (nsps) involving viral replication and transcription. Most prominently, nsp12 contains RNA-dependent RNA polymerase (RdRP) which catalyzes the template-directed viral RNA synthesis.^12^ Thus, RdRP^13-17^, 3CLpro^18-20^, and host-viral fusion^21, 22^ are promising targets for antiviral agents against SARS-CoV-2. In line with the knowledge that the severity of COVID-19 is strongly associated with viral replication level^23, 24^, it has been hypothesized that the direct-acting antiviral agents will be an effective approach in alleviating aggravation of COVID-19 symptoms and preventing the further spread of COVID-19.

Clevudine is an antiviral drug to treat chronic hepatitis B virus (HBV) infection. Unlike SARS-CoV-2, HBV has a relaxed-circular partially double DNA (rcDNA) genome. A covalently closed circular DNA (cccDNA)--the viral intermediate formed by DNA repair process in rcDNA--is transcribed into pregenomic RNA (pgRNA) by host DNA-dependent RNA polymerase. Thereafter, reverse transcription of pgRNA by RNA-dependent DNA polymerase generates rcDNA for viral shedding.^25^ Although clevudine is a pyrimidine analogue, it has antiviral effects on HBV by inhibiting the DNA polymerization of protein priming rather than incorporating the viral DNA.^26, 27^ Based on this action mode of clevudine, it is expected that clevudine would have antiviral effects on SARS-CoV-2. Furthermore, recent molecular docking studies revealed that clevudine has strong inhibitory capabilities against 3CLpro and ACE2 of SARS-CoV-2.^28, 29^ Altogether, clevudine is hypothesized to inhibit the replication of SARS-CoV-2 virus. Therefore, we conducted a prospective, single-blind, proof of concept clinical study to examine the antiviral efficacy and safety of clevudine compared to placebo in Korean COVID-19 patients with moderate severity (BK-CLV-201, ClinicalTrials.gov identifier NCT04347915).

## Methods

### Clinical study oversight

Adults aged 19 years or older were eligible if they showed real-time reverse transcription polymerase chain reaction (RT-PCR) positive result for SARS-CoV-2 infection within 96 hours and had symptoms of COVID-19 within 7 days from screening to treatment initiation. Participants were randomized 2:1 to 120 mg clevudine or matching placebo to receive one of treatments orally once-daily for 14 days (Figure 1). Planned number of patients was 60 and this sample size was chosen arbitrarily. Antiviral efficacy and safety were evaluated for 29 days after treatment initiation. For assessment of antiviral efficacy, nasopharyngeal swabs were collected at Day 1 (baseline), 4, 8, 11, 15, 22 and 29 for real-time RT-PCR test. Clinical laboratory tests were evaluated at Day 1, 4, 8, 15, 22 and 29, and adverse events (AEs) were monitored throughout the study.

**Figure 1.**
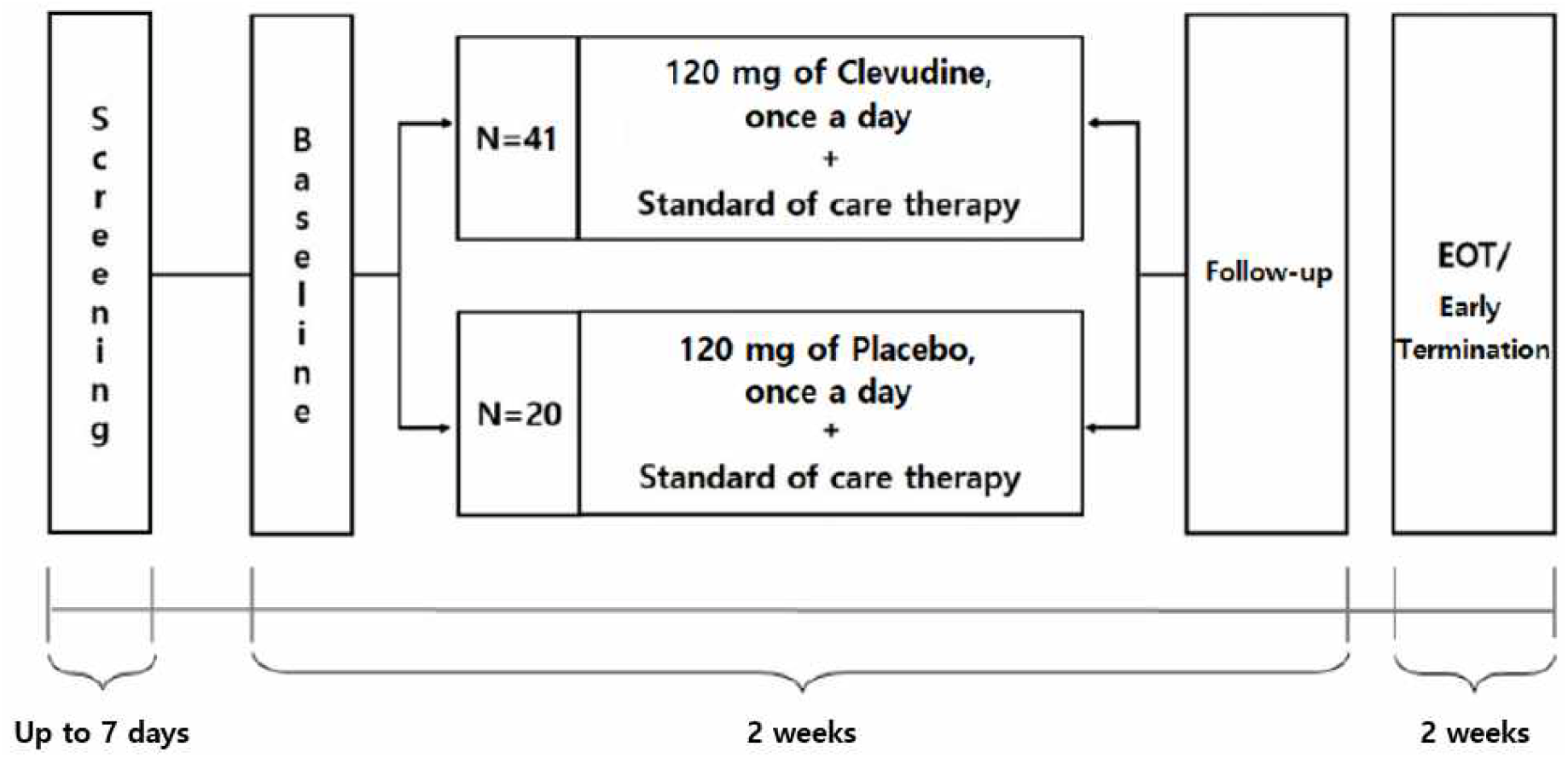
Clinical study oversight.

### Antiviral efficacy outcome

The primary antiviral efficacy outcome was the proportion of patients with real-time RT-PCR negative result for SARS-CoV-2 infection at Day 15. Secondary antiviral efficacy outcomes were the proportion of patients with real-time RT-PCR negative result at Day 4, 8, 11, 22 and 29; as well as the changes in cycle threshold (Ct) value of E, RdRP, and N genes from baseline to Day 4, 8, 11, 15, 22 and 29. Real-time RT-PCR test for SARS-CoV-2 infection was performed using Allplex^TM^ 2019-nCoV Assay kit and sample was considered negative if all Ct values of target genes (E, RdRP, and N genes) were 40 or higher.

### Clinical efficacy outcome

Clinical efficacy outcomes included proportion of patients who showed improvement in lung involvement by imaging tests (e.g. X-Ray, CT-Scan) from baseline to Day 29, proportion of patients with normal body temperature, proportion of patients with normal oxygen saturation (SpO2, ≥ 95%) at Day 4, 8, 11, 15, 22 and 29, and the changes in C-reactive protein (CRP) from baseline to Day 4, 8, 15, 22 and 29.

### Safety outcome

Safety outcomes included changes in clinical laboratory tests, vital signs measurement, and physical examination from baseline, and incidence of AEs and serious AEs (SAEs).

### Statistical analysis

Efficacy analyses were conducted in intent-to-treat (ITT) population. The number and percentage of patients with real-time RT-PCR negative result for SARS-CoV-2 infection, patients with normal body temperature, patients with normal oxygen saturation at Day 4, 8, 11, 15, 22 and 29, and patients who showed improvement in lung involvement at Day 29 were summarized and between-treatment comparisons were conducted using Pearson’s chi-square test or Fisher’s Exact test. Changes in Ct value of E, RdRP, N genes, and CRP from baseline were presented as descriptive statistics (i.e. mean, standard deviations, median, min, max) and comparisons between treatments at each visit were conducted using a Two-sample t-test or Wilcoxon’s rank sum test. Analyses were performed using SAS Version 9.4 (SAS Institute Inc., Cary NC) and two-sided tests were performed using an alpha of 0.1 for between-treatment comparisons. Safety analyses were based on the safety population which consisted of all patients who received at least 1 dose of study drug. The number of patients with AEs and adverse drug reactions (ADRs) in each treatment group were summarized using frequency distribution. Test values and change from baseline were summarized descriptively by treatment group for clinical laboratory test results, vital signs, and physical examination.

## Results

### Patient Characteristics

A total of 61 patients were analyzed as ITT population and safety population of which 41 and 20 patients were randomized in clevudine and placebo group, respectively. The mean age at baseline was 60.02 (±13.44) and 59.50 (±11.69) years in clevudine and placebo group, respectively. The most common clinical symptom of COVID-19 among all patients were fever (90.16%), cough (65.57%), and myalgia (44.26%). A total of 28 patients (45.90%) had hypertension at baseline, of which 19 and 9 patients were randomized in clevudine and placebo group, respectively. Of all patients, 34 and 49 patients were randomized within 5 and 7 days from the symptom onset, respectively. Baseline characteristics of the patients are shown in Table 1.

**Table 1.** Baseline characteristics of the patients.

|  |  | Clevudine<br>(N=41) | Placebo<br>(N=20) | Total<br>(N=61) |
| --- | --- | --- | --- | --- |
| Age | Mean±Std | 60.02±13.44 | 59.50±11.69 | 59.85±12.80 |
|  | Median | 63.00 | 60.00 | 61.00 |
|  | Min~Max | 24.00~81.00 | 36.00~74.00 | 24.00~81.00 |
| Gender | Male | 20 (48.78%) | 10 (50.00%) | 30 (49.18%) |
|  | Female | 21 (51.22%) | 10 (50.00%) | 31 (50.82%) |
| Symptoms of COVID-19 | Fever | 39 (95.12%) | 16 (80.00%) | 55 (90.16%) |
|  | Cough | 23 (56.10%) | 17 (85.00%) | 40 (65.57%) |
|  | Myalgia | 19 (46.34%) | 8 (40.00%) | 27 (44.26%) |
|  | Headache | 16 (39.02%) | 11 (55.00%) | 27 (44.26%) |
|  | Sore throat | 15 (36.59%) | 9 (45.00%) | 24 (39.34%) |
| Day from symptom onset to<br>randomization | 1~5 | 21 (51.22%) | 13 (65.00%) | 34 (55.74%) |
|  | 1~7 | 32 (78.05%) | 17 (85.00%) | 49 (80.33%) |
| Comorbidity | Hypertension | 19 (46.34%) | 9 (45.00%) | 28 (45.90%) |
|  | Diabetes mellitus | 13 (31.71%) | 3 (15.00%) | 16 (26.23%) |
|  | Hyperlipidemia | 5 (12.20%) | 3 (15.00%) | 8 (13.11%) |

### Proportion of patients with real-time RT-PCR negative test result

We evaluated the proportion of patients with real-time RT-PCR negative test result at Day 4, 8, 11, 15, 22, and 29. 2 patients of ITT population were excluded from the analysis because they had negative result at baseline. There was no significant difference in the proportion of patients with negative result between clevudine group and placebo group at every visit (Table 2).

**Table 2.**
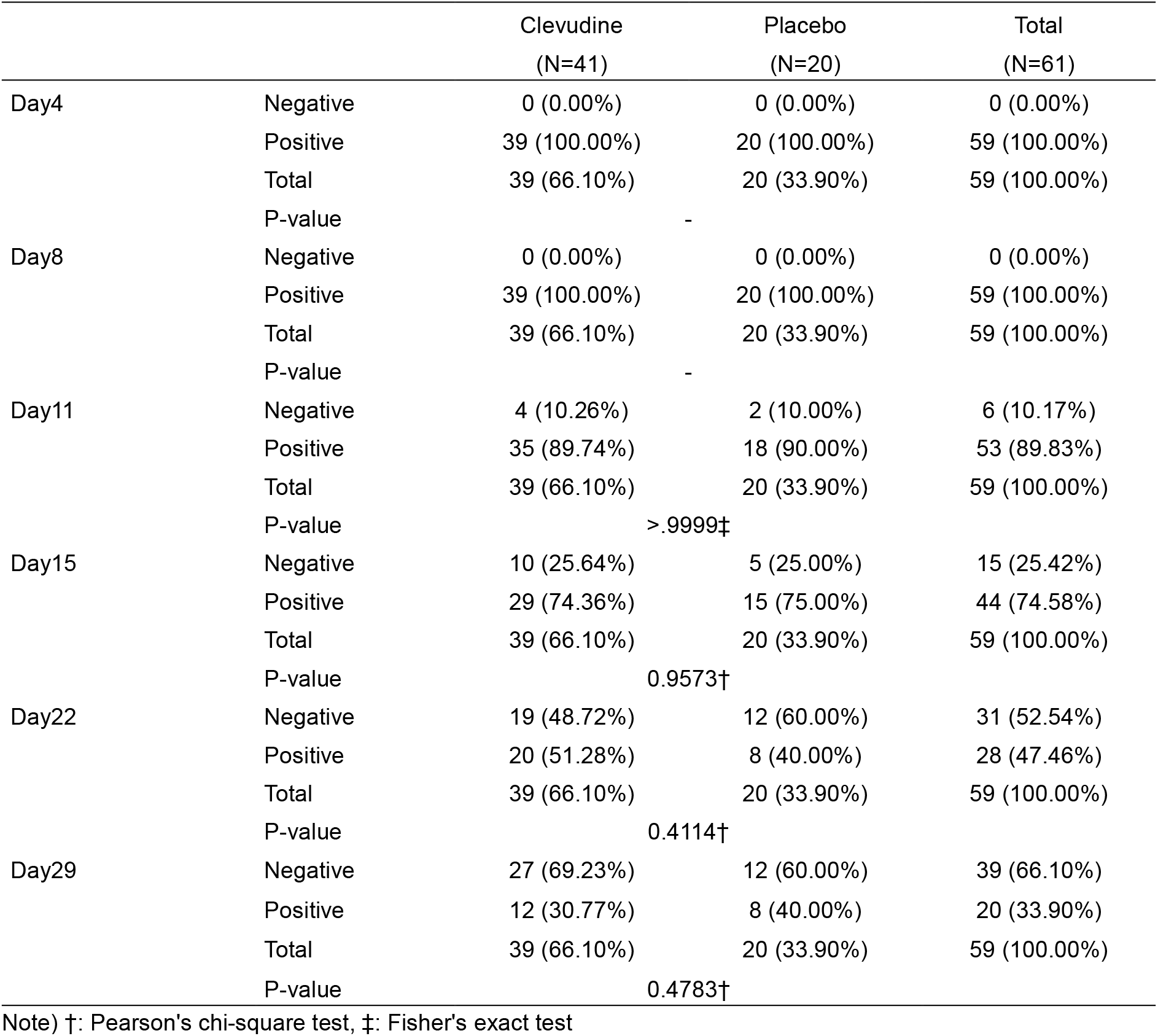
Proportion of patients with real-time RT-PCR negative test result (ITT set)

|  |  | Clevudine<br>(N=41) | Placebo<br>(N=20) | Total<br>(N=61) |
| --- | --- | --- | --- | --- |
| Day4 | Negative | 0 (0.00%) | 0 (0.00%) | 0 (0.00%) |
|  | Positive | 39 (100.00%) | 20 (100.00%) | 59 (100.00%) |
|  | Total | 39 (66.10%) | 20 (33.90%) | 59 (100.00%) |
|  | P-value | - |  |  |
| Day8 | Negative | 0 (0.00%) | 0 (0.00%) | 0 (0.00%) |
|  | Positive | 39 (100.00%) | 20 (100.00%) | 59 (100.00%) |
|  | Total | 39 (66.10%) | 20 (33.90%) | 59 (100.00%) |
|  | P-value | - |  |  |
| Day11 | Negative | 4 (10.26%) | 2 (10.00%) | 6 (10.17%) |
|  | Positive | 35 (89.74%) | 18 (90.00%) | 53 (89.83%) |
|  | Total | 39 (66.10%) | 20 (33.90%) | 59 (100.00%) |
|  | P-value | >.9999‡ |  |  |
| Day15 | Negative | 10 (25.64%) | 5 (25.00%) | 15 (25.42%) |
|  | Positive | 29 (74.36%) | 15 (75.00%) | 44 (74.58%) |
|  | Total | 39 (66.10%) | 20 (33.90%) | 59 (100.00%) |
|  | P-value | 0.9573† |  |  |
| Day22 | Negative | 19 (48.72%) | 12 (60.00%) | 31 (52.54%) |
|  | Positive | 20 (51.28%) | 8 (40.00%) | 28 (47.46%) |
|  | Total | 39 (66.10%) | 20 (33.90%) | 59 (100.00%) |
|  | P-value | 0.4114† |  |  |
| Day29 | Negative | 27 (69.23%) | 12 (60.00%) | 39 (66.10%) |
|  | Positive | 12 (30.77%) | 8 (40.00%) | 20 (33.90%) |
|  | Total | 39 (66.10%) | 20 (33.90%) | 59 (100.00%) |
|  | P-value | 0.4783† |  |  |

Note) †: Pearson's chi-square test, ‡: Fisher's exact test

### Changes in Ct value of E, RdRP, N genes from baseline

Changes in Ct values of E, RdRP, and N genes from baseline to Day 4, 8, 11, 15, 22, and 29 were evaluated. Ct values provided by real-time RT-PCR are highly associated with the copy number of viral RNA and viral load, implicating that higher Ct values relate to lower viral loads.^30, 31^ Ct values of all genes at baseline were significantly lower in clevudine group compared to placebo group. Considering that lower Ct values are related to a higher amount of target viral sequence, there was an imbalance in viral load between treatment groups. Clevudine group showed greater change in Ct values of all genes from baseline to all visits (except Day 8) than placebo group, but there was no significant difference between clevudine group and placebo group at every visit (Table 3 and Figure 2).

**Figure 2.**
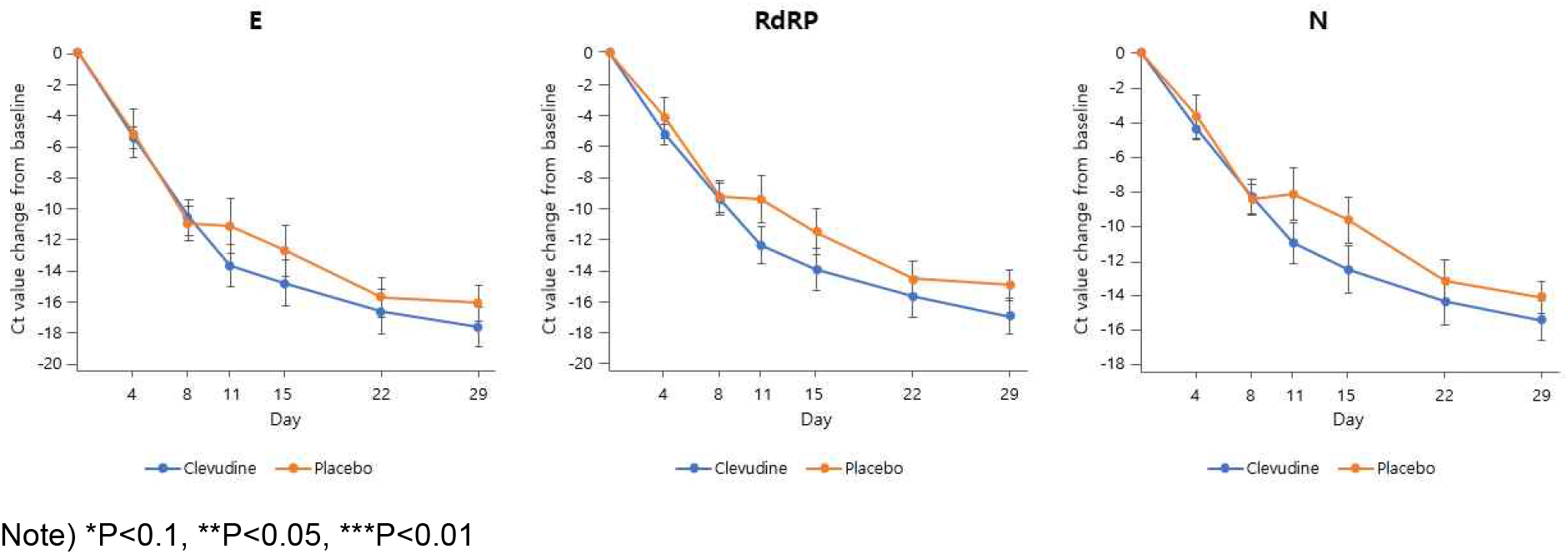
Change in Ct value from baseline (ITT set)

**Table 3.** Change in Ct value from baseline (ITT set)

|  |  | E gene |  | RdRP gene |  | N gene |  |
| --- | --- | --- | --- | --- | --- | --- | --- |
|  |  | Clevudine<br>(N=41) | Placebo<br>(N=20) | Clevudine<br>(N=41) | Placebo<br>(N=20) | Clevudine<br>(N=41) | Placebo<br>(N=20) |
| Baseline | N | 39 | 20 | 39 | 20 | 39 | 20 |
|  | Mean±Std | 21.57±6.76 | 24.72±6.51 | 22.45±6.29 | 25.35±5.55 | 23.55±6.29 | 25.96±5.27 |
|  | Median | 21.05 | 23.09 | 21.91 | 24.55 | 23.27 | 25.71 |
|  | Min~Max | 11.07~40.00 | 11.87~40.00 | 13.01~36.45 | 13.65~37.05 | 14.57~40.00 | 11.91~35.84 |
|  | P-value | 0.0928* |  | 0.0862* |  | 0.1469* |  |
| Day4 | N | 39 | 20 | 39 | 20 | 39 | 20 |
|  | Mean±Std | 27.02±6.68 | 29.89±7.05 | 27.66±6.03 | 29.52±5.59 | 27.91±5.22 | 29.64±5.05 |
|  | Median | 27.03 | 27.20 | 28.08 | 28.29 | 28.38 | 29.02 |
|  | Min~Max | 15.38~40.00 | 20.72~40.00 | 15.26~40.00 | 20.26~40.00 | 15.55~35.81 | 20.36~38.83 |
|  | P-value | 0.2436 ¥ |  | 0.2556* |  | 0.2271* |  |
| Day8 | N | 36 | 18 | 36 | 18 | 36 | 18 |
|  | Mean±Std | 32.35±6.50 | 35.05±5.84 | 32.02±5.24 | 34.16±5.03 | 32.04±4.59 | 34.04±4.77 |
|  | Median | 32.94 | 38.23 | 33.62 | 34.79 | 33.32 | 34.67 |
|  | Min~Max | 19.28~40.00 | 24.69~40.00 | 20.28~40.00 | 23.31~40.00 | 22.68~40.00 | 24.00~40.00 |
|  | P-value | 0.1766 ¥ |  | 0.1572* |  | 0.1422* |  |
| Day11 | N | 31 | 18 | 31 | 18 | 31 | 18 |
|  | Mean±Std | 35.95±5.38 | 35.22±4.87 | 35.35±4.61 | 34.30±4.36 | 35.15±4.12 | 33.76±3.88 |
|  | Median | 40.00 | 34.98 | 36.51 | 33.46 | 35.42 | 33.33 |
|  | Min~Max | 24.17~40.00 | 25.76~40.00 | 25.67~40.00 | 26.71~40.00 | 26.40~40.00 | 27.95~40.00 |
|  | P-value | 0.5997 ¥ |  | 0.4144 ¥ |  | 0.2296 ¥ |  |
| Day15 | N | 34 | 18 | 34 | 18 | 34 | 18 |
|  | Mean±Std | 36.73±5.47 | 36.81±4.60 | 36.76±4.92 | 36.41±4.28 | 36.49±4.61 | 35.24±4.28 |
|  | Median | 40.00 | 40.00 | 40.00 | 38.55 | 37.46 | 35.33 |
|  | Min~Max | 16.29~40.00 | 24.90~40.00 | 18.13~40.00 | 26.91~40.00 | 19.31~40.00 | 26.24~40.00 |
|  | P-value | 0.8784 ¥ |  | 0.6501 ¥ |  | 0.1983 ¥ |  |
|  |  | Clevudine<br>(N=41) | Placebo<br>(N=20) | Clevudine<br>(N=41) | Placebo<br>(N=20) | Clevudine<br>(N=41) | Placebo<br>(N=20) |
| Day22 | N | 30 | 18 | 30 | 18 | 30 | 18 |
|  | Mean±Std | 38.69±2.78 | 39.82±0.76 | 38.60±2.75 | 39.44±1.39 | 38.48±2.60 | 38.77±2.33 |
|  | Median | 40.00 | 40.00 | 40.00 | 40.00 | 40.00 | 40.00 |
|  | Min~Max | 31.40~40.00 | 36.76~40.00 | 31.69~40.00 | 35.69~40.00 | 32.35~40.00 | 31.76~40.00 |
|  | P-value | 0.1566 ¥ |  | 0.4751 ¥ |  | 0.7920 ¥ |  |
| Day29 | N | 32 | 17 | 32 | 17 | 32 | 17 |
|  | Mean±Std | 39.30±2.24 | 39.66±1.42 | 39.47±2.07 | 39.43±1.80 | 39.20±2.35 | 39.32±1.85 |
|  | Median | 40.00 | 40.00 | 40.00 | 40.00 | 40.00 | 40.00 |
|  | Min~Max | 31.43~40.00 | 34.16~40.00 | 31.50~40.00 | 32.91~40.00 | 30.77~40.00 | 33.42~40.00 |
|  | P-value | 0.6444 ¥ |  | 0.5829 ¥ |  | 0.6932 ¥ |  |
| Day4 - Baseline<br>(Change) | N | 39 | 20 | 39 | 20 | 39 | 20 |
|  | Mean±Std | 5.45±4.43 | 5.18±7.01 | 5.22±4.04 | 4.17±5.81 | 4.36±4.28 | 3.68±5.58 |
|  | Median | 5.57 | 4.90 | 5.87 | 5.15 | 4.50 | 3.99 |
|  | Min~Max | -4.74~16.48 | -17.13~17.33 | -4.92~14.66 | -14.99~12.89 | -6.01~13.79 | -14.68~13.09 |
| | P-value (within) | <.0001# | 0.0009\$ | <.0001# | 0.0017\$ | <.0001# | 0.0012\$ |
|  | P-value (between) | 0.9619 ¥ |  | 0.4405 ¥ |  | 0.6613 ¥ |  |
| Day8 - Baseline<br>(Change) | N | 36 | 18 | 36 | 18 | 36 | 18 |
|  | Mean±Std | 10.60±6.87 | 10.97±4.71 | 9.43±6.18 | 9.27±4.40 | 8.34±6.13 | 8.48±3.60 |
|  | Median | 10.88 | 12.62 | 9.31 | 9.09 | 8.51 | 8.50 |
|  | Min~Max | -1.78~24.34 | 3.08~17.33 | -1.72~21.02 | 3.27~16.24 | -6.72~19.12 | 3.86~14.26 |
|  | P-value (within) | <.0001# | <.0001# | <.0001# | <.0001# | <.0001# | <.0001# |
|  | P-value (between) | 0.8378* |  | 0.9225* |  | 0.9188* |  |
|  |  | Clevudine<br>(N=41) | Placebo<br>(N=20) | Clevudine<br>(N=41) | Placebo<br>(N=20) | Clevudine<br>(N=41) | Placebo<br>(N=20) |
| Day11 - Baseline | N | 31 | 18 | 31 | 18 | 31 | 18 |
| (Change) | Mean±Std | 13.70±7.57 | 11.14±7.50 | 12.39±6.68 | 9.41±6.60 | 10.99±6.59 | 8.19±6.44 |
|  | Median | 12.40 | 12.18 | 11.31 | 9.08 | 9.85 | 7.49 |
|  | Min~Max | 0.00~27.74 | -2.69~28.13 | 0.23~25.97 | -2.56~19.06 | -5.21~24.27 | -3.33~22.01 |
|  | P-value (within) | <.0001# | <.0001# | <.0001# | <.0001# | <.0001# | <.0001# |
|  | P-value (between) | 0.2568* |  | 0.1386* |  | 0.1541* |  |
| Day15 - Baseline | N | 34 | 18 | 34 | 18 | 34 | 18 |
| (Change) | Mean±Std | 14.83±8.67 | 12.73±6.88 | 13.95±7.86 | 11.52±6.24 | 12.55±7.89 | 9.67±5.63 |
|  | Median | 15.02 | 13.62 | 13.44 | 12.44 | 12.59 | 9.52 |
|  | Min~Max | -5.32~28.93 | -1.59~24.06 | -4.49~25.60 | -1.03~22.84 | -5.95~24.63 | -1.95~20.15 |
|  | P-value (within) | <.0001# | <.0001# | <.0001# | <.0001# | <.0001# | <.0001# |
|  | P-value (between) | 0.3784* |  | 0.2612* |  | 0.1765* |  |
| Day22 - Baseline | N | 30 | 18 | 30 | 18 | 30 | 18 |
| (Change) | Mean±Std | 16.65±7.73 | 15.74±5.32 | 15.69±7.18 | 14.55±4.98 | 14.39±7.35 | 13.20±5.06 |
|  | Median | 16.75 | 16.91 | 16.45 | 15.46 | 14.50 | 13.77 |
|  | Min~Max | 0.00~28.16 | 5.08~24.89 | 1.21~25.97 | 3.79~22.84 | -4.18~24.63 | 0.30~23.12 |
|  | P-value (within) | <.0001# | <.0001# | <.0001# | <.0001# | <.0001# | <.0001# |
|  | P-value (between) | 0.6623* |  | 0.5563* |  | 0.5472* |  |
| Day29 - Baseline | N | 32 | 17 | 32 | 17 | 32 | 17 |
| (Change) | Mean±Std | 17.63±7.27 | 16.11±4.73 | 16.96±6.65 | 14.95±4.06 | 15.51±6.69 | 14.16±3.80 |
|  | Median | 16.22 | 16.96 | 16.79 | 15.71 | 13.84 | 14.26 |
|  | Min~Max | 0.00~28.93 | 5.08~24.06 | 3.55~26.99 | 5.75~22.84 | 0.00~25.43 | 7.17~21.67 |
|  | P-value (within) | <.0001# | <.0001# | <.0001# | <.0001# | <.0001# | <.0001# |
|  | P-value (between) | 0.4433* |  | 0.1973* |  | 0.3729* |  |

Note) \*: Two sample t-test, ¥ : Wilcoxon's rank sum test, #: Paired t-test, \$: Wilcoxon's signed rank test

We conducted subgroup analysis by hypertension and time from the symptom onset to randomization on change in Ct values of E, RdRP, and N genes from baseline.

In patients with hypertension, the changes in Ct values of E, RdRP, and N genes from baseline to Day 15, 22, and 29 were significantly greater in clevudine group compared to placebo group (Table 4 and Figure 3; E gene: Day 15: p=0.0306, Day 22: p=0.0543, Day 29: p=0.0367; RdRP gene: Day 15: p=0.0249, Day 22: p=0.0435, Day 29: p=0.0111; N gene: Day 15: p=0.0212, Day 22: p=0.0369, Day 29: p=0.0308). Furthermore, the changes in Ct values of E, RdRP, and N genes from baseline to Day 15, 22, and 29 were significantly greater in clevudine group compared to placebo group in patients with hypertension and randomized within the first 7 days after the onset of symptoms (Table 5 and Figure 4; E gene; Day 15 p=0.0080, Day 22 p=0.0295, Day 29 p=0.0577; RdRP gene: Day 15 p=0.0054, Day 22 p=0.0202, Day 29 p=0.0185; N gene: Day 15 p=0.0082, Day 22 p=0.0187, Day 29 p=0.0464).

**Figure 3.**
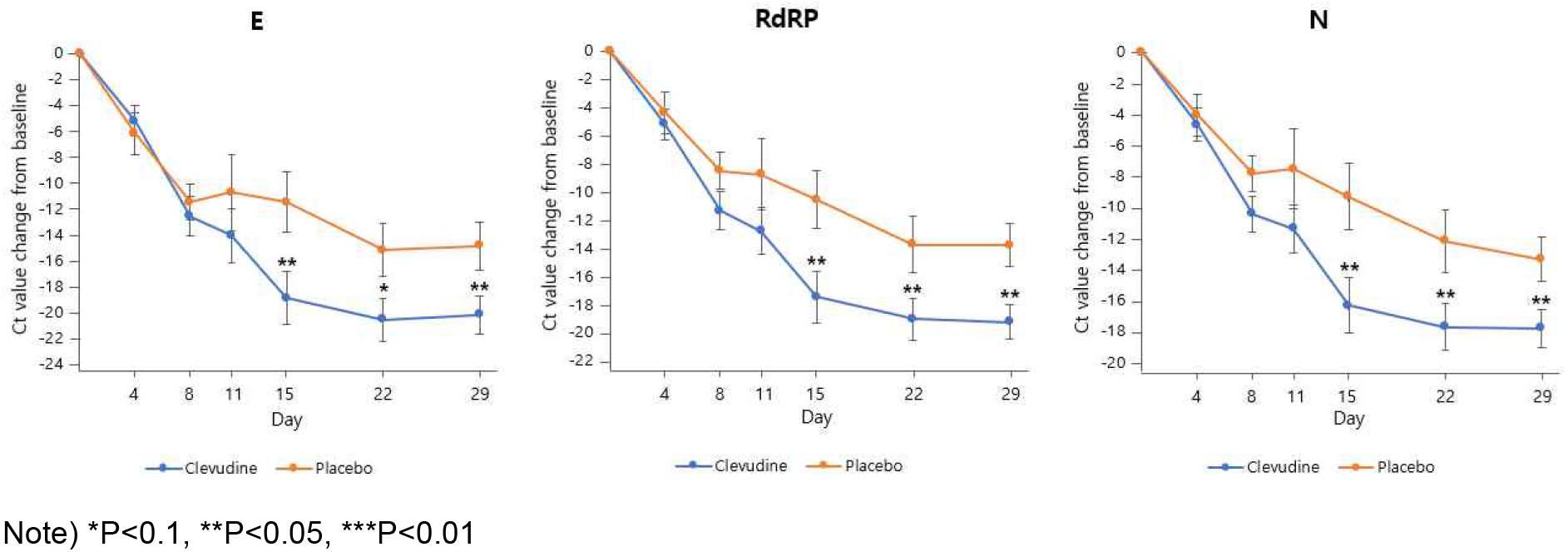
Change in Ct value from baseline (patients with hypertension, ITT set)

**Figure 4.**
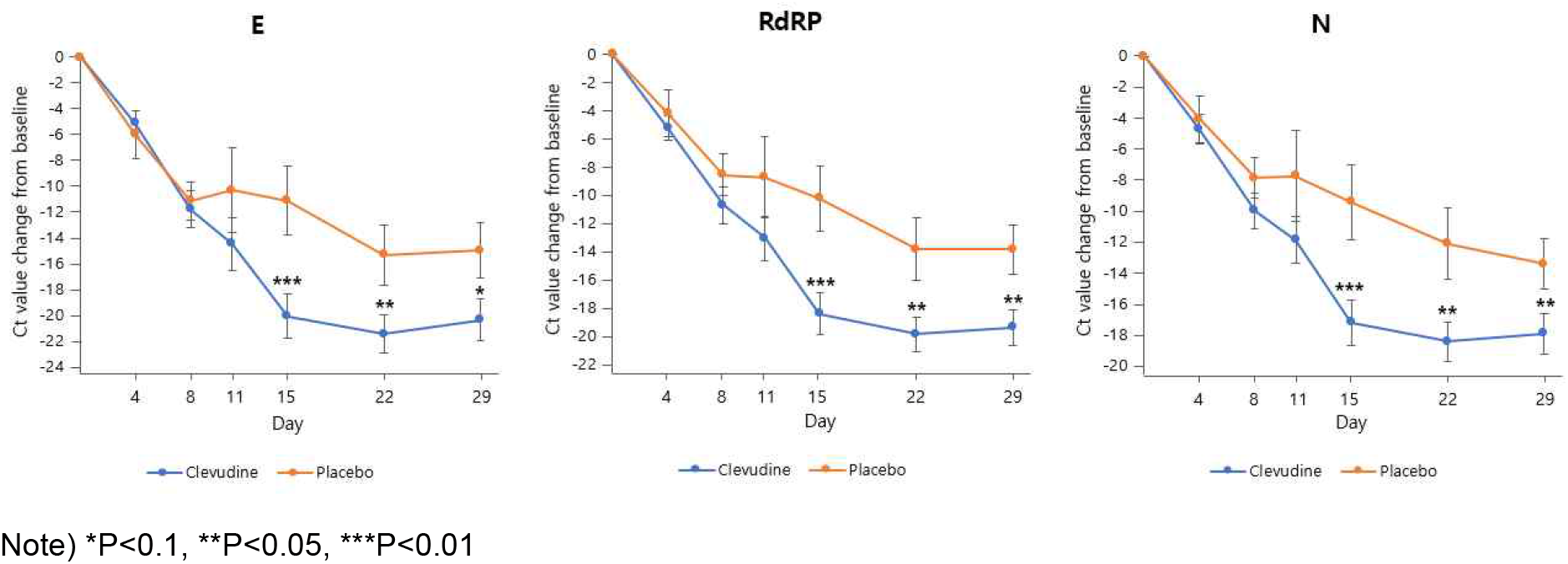
Change in Ct value from baseline (patients with hypertension and randomized within 7 days after symptom onset, ITT set)

**Table 4.** Change in Ct value from baseline (patients with hypertension, ITT set)

|  |  | E gene |  | RdRP gene |  | N gene |  |
| --- | --- | --- | --- | --- | --- | --- | --- |
|  |  | Clevudine<br>(N=19) | Placebo<br>(N=9) | Clevudine<br>(N=19) | Placebo<br>(N=9) | Clevudine<br>(N=19) | Placebo<br>(N=9) |
| Baseline | N | 19 | 9 | 19 | 9 | 19 | 9 |
|  | Mean±Std | 18.66±4.31 | 24.45±6.86 | 19.83±3.70 | 25.13±6.17 | 20.88±3.61 | 25.37±6.12 |
|  | Median | 19.63 | 23.14 | 20.18 | 24.83 | 20.80 | 26.39 |
|  | Min~Max | 11.84~25.35 | 11.87~34.92 | 14.40~26.38 | 13.65~34.25 | 15.37~27.54 | 11.91~32.83 |
|  | P-value | 0.0110* |  | 0.0085* |  | 0.0214* |  |
| Day4 | N | 19 | 9 | 19 | 9 | 19 | 9 |
|  | Mean±Std | 23.89±5.18 | 30.64±7.87 | 25.05±4.99 | 29.51±6.29 | 25.56±4.74 | 29.42±5.37 |
|  | Median | 24.50 | 33.25 | 24.45 | 32.77 | 25.33 | 31.14 |
|  | Min~Max | 15.38~32.14 | 20.72~40.00 | 15.26~31.99 | 20.26~36.45 | 15.55~32.50 | 20.36~35.13 |
|  | P-value | 0.0115* |  | 0.0521* |  | 0.0649* |  |
| Day8 | N | 17 | 9 | 17 | 9 | 17 | 9 |
|  | Mean±Std | 30.96±6.59 | 35.93±6.00 | 30.86±5.51 | 33.65±5.62 | 31.07±4.85 | 33.18±4.55 |
|  | Median | 31.12 | 40.00 | 31.65 | 34.86 | 32.58 | 34.74 |
|  | Min~Max | 19.28~40.00 | 24.69~40.00 | 20.28~40.00 | 23.31~40.00 | 22.68~40.00 | 24.00~37.42 |
|  | P-value | 0.0952 ¥ |  | 0.2357* |  | 0.2684 ¥ |  |
| Day11 | N | 13 | 9 | 13 | 9 | 13 | 9 |
|  | Mean±Std | 33.17±5.95 | 35.21±4.74 | 32.87±4.87 | 33.89±4.30 | 32.68±4.07 | 32.91±4.05 |
|  | Median | 32.48 | 34.26 | 33.52 | 32.71 | 32.87 | 33.27 |
|  | Min~Max | 24.17~40.00 | 29.30~40.00 | 25.67~40.00 | 28.49~40.00 | 26.40~40.00 | 28.13~40.00 |
|  | P-value | 0.4208 ¥ |  | 0.6202* |  | 0.8995* |  |
| Day15 | N | 15 | 9 | 15 | 9 | 15 | 9 |
|  | Mean±Std | 37.21±4.68 | 35.93±4.07 | 37.07±4.06 | 35.70±4.28 | 37.08±3.81 | 34.69±4.40 |
|  | Median | 40.00 | 35.89 | 38.80 | 35.42 | 37.36 | 33.97 |
|  | Min~Max | 24.49~40.00 | 31.37~40.00 | 25.82~40.00 | 30.87~40.00 | 26.80~40.00 | 29.51~40.00 |
|  | P-value | 0.3610 ¥ |  | 0.4234 ¥ |  | 0.1964 ¥ |  |
|  |  | Clevudine<br>(N=19) | Placebo<br>(N=9) | Clevudine<br>(N=19) | Placebo<br>(N=9) | Clevudine<br>(N=19) | Placebo<br>(N=9) |
| Day22 | N | 14 | 9 | 14 | 9 | 14 | 9 |
|  | Mean±Std | 38.66±2.77 | 39.64±1.08 | 38.61±2.89 | 38.89±1.85 | 38.45±2.75 | 37.54±2.85 |
|  | Median | 40.00 | 40.00 | 40.00 | 40.00 | 40.00 | 37.46 |
|  | Min~Max | 31.63~40.00 | 36.76~40.00 | 31.69~40.00 | 35.69~40.00 | 32.35~40.00 | 31.76~40.00 |
|  | P-value | 0.4822 ¥ |  | 0.7172 ¥ |  | 0.3108 ¥ |  |
| Day29 | N | 15 | 9 | 15 | 9 | 15 | 9 |
|  | Mean±Std | 38.50±3.14 | 39.35±1.95 | 38.88±2.96 | 38.92±2.41 | 38.57±3.20 | 38.72±2.45 |
|  | Median | 40.00 | 40.00 | 40.00 | 40.00 | 40.00 | 40.00 |
|  | Min~Max | 31.43~40.00 | 34.16~40.00 | 31.50~40.00 | 32.91~40.00 | 30.77~40.00 | 33.42~40.00 |
|  | P-value | 0.5268 ¥ |  | 0.7508 ¥ |  | 0.6426 ¥ |  |
| Day4 - Baseline<br>(Change) | N | 19 | 9 | 19 | 9 | 19 | 9 |
|  | Mean±Std | 5.22±5.12 | 6.19±4.91 | 5.22±4.63 | 4.38±4.43 | 4.68±4.71 | 4.05±4.04 |
|  | Median | 5.49 | 6.90 | 5.81 | 5.55 | 4.96 | 3.91 |
|  | Min~Max | -4.74~16.48 | -1.58~13.11 | -4.92~14.66 | -2.90~12.89 | -5.52~13.79 | -2.18~11.46 |
|  | P-value (within) | 0.0003# | 0.0054# | 0.0001# | 0.0180# | 0.0004# | 0.0169# |
|  | P-value (between) | 0.6414* |  | 0.6552* |  | 0.7318* |  |
| Day8 - Baseline<br>(Change) | N | 17 | 9 | 17 | 9 | 17 | 9 |
|  | Mean±Std | 12.58±6.13 | 11.48±4.10 | 11.34±5.44 | 8.51±3.95 | 10.44±4.71 | 7.81±3.39 |
|  | Median | 12.93 | 12.82 | 10.84 | 8.37 | 10.70 | 7.51 |
|  | Min~Max | -1.45~24.34 | 5.08~16.86 | -1.63~19.76 | 4.00~15.74 | -0.59~18.04 | 3.97~12.09 |
|  | P-value (within) | <.0001# | <.0001# | <.0001# | 0.0002# | <.0001# | 0.0001# |
|  | P-value (between) | 0.6339* |  | 0.1824* |  | 0.1530* |  |
|  |  | Clevudine | Placebo | Clevudine | Placebo | Clevudine | Placebo |
|  |  | (N=19) | (N=9) | (N=19) | (N=9) | (N=19) | (N=9) |
| Day11 - Baseline | N | 13 | 9 | 13 | 9 | 13 | 9 |
| (Change) | Mean±Std | 14.08±7.63 | 10.76±8.83 | 12.77±6.09 | 8.76±7.51 | 11.39±5.48 | 7.54±7.71 |
|  | Median | 13.51 | 9.21 | 11.63 | 9.08 | 10.48 | 6.04 |
|  | Min~Max | 5.18~27.74 | -2.69~28.13 | 4.91~23.32 | -2.56~19.06 | 3.98~20.57 | -3.33~22.01 |
|  | P-value (within) | <.0001# | 0.0064# | <.0001# | 0.0081# | <.0001# | 0.0189# |
|  | P-value (between) | 0.3571* |  | 0.1823* |  | 0.1849* |  |
| Day15 - Baseline | N | 15 | 9 | 15 | 9 | 15 | 9 |
| (Change) | Mean±Std | 18.93±7.96 | 11.48±7.08 | 17.45±7.07 | 10.56±6.24 | 16.33±6.86 | 9.32±6.41 |
|  | Median | 21.18 | 11.11 | 19.40 | 10.13 | 18.83 | 7.53 |
|  | Min~Max | -0.53~28.16 | -1.59~20.69 | -0.56~25.60 | -1.03~18.38 | -0.74~24.63 | -1.95~20.15 |
|  | P-value (within) | <.0001# | 0.0012# | <.0001# | 0.0010# | <.0001# | 0.0024# |
|  | P-value (between) | 0.0306* |  | 0.0249* |  | 0.0212* |  |
| Day22 - Baseline | N | 14 | 9 | 14 | 9 | 14 | 9 |
| (Change) | Mean±Std | 20.58±6.21 | 15.19±6.16 | 19.04±5.64 | 13.75±5.95 | 17.70±5.62 | 12.17±6.10 |
|  | Median | 21.28 | 16.86 | 19.45 | 15.17 | 17.97 | 12.77 |
|  | Min~Max | 6.61~28.16 | 5.08~24.89 | 5.31~25.60 | 3.79~22.13 | 4.81~24.63 | 0.30~23.12 |
|  | P-value (within) | <.0001# | <.0001# | <.0001# | 0.0001# | <.0001# | 0.0003# |
|  | P-value (between) | 0.0543* |  | 0.0435* |  | 0.0369* |  |
| Day29 - Baseline | N | 15 | 9 | 15 | 9 | 15 | 9 |
| (Change) | Mean±Std | 20.22±5.67 | 14.90±5.69 | 19.25±4.71 | 13.79±4.62 | 17.81±4.72 | 13.35±4.33 |
|  | Median | 19.27 | 16.86 | 19.24 | 15.17 | 16.73 | 12.77 |
|  | Min~Max | 11.30~28.16 | 5.08~22.29 | 11.21~25.60 | 5.75~19.26 | 10.51~24.63 | 7.17~21.51 |
|  | P-value (within) | <.0001# | <.0001# | <.0001# | <.0001# | <.0001# | <.0001# |
|  | P-value (between) | 0.0367* |  | 0.0111* |  | 0.0308* |  |

Note) \*: Two sample t-test, ¥ : Wilcoxon's rank sum test, #: Paired t-test

**Table 5.** Change in Ct value from baseline (patients with hypertension and randomized within 7 days after symptom onset, ITT set)

|  |  | E gene |  | RdRP gene |  | N gene |  |
| --- | --- | --- | --- | --- | --- | --- | --- |
|  |  | Clevudine<br>(N=16) | Placebo<br>(N=8) | Clevudine<br>(N=16) | Placebo<br>(N=8) | Clevudine<br>(N=16) | Placebo<br>(N=8) |
| Baseline | N | 16 | 8 | 16 | 8 | 16 | 8 |
|  | Mean±Std | 18.31±4.30 | 24.33±7.32 | 19.59±3.59 | 24.93±6.57 | 20.63±3.47 | 25.14±6.50 |
|  | Median | 18.59 | 23.075 | 19.815 | 23.685 | 20.78 | 25.34 |
|  | Min~Max | 11.84~25.35 | 11.87~34.92 | 14.40~26.29 | 13.65~34.25 | 15.37~26.79 | 11.91~32.83 |
|  | P-value | 0.0182* |  | 0.0598* |  | 0.0994* |  |
| Day4 | N | 16 | 8 | 16 | 8 | 16 | 8 |
|  | Mean±Std | 23.44±4.54 | 30.32±8.35 | 24.82±4.18 | 29.11±6.60 | 25.36±3.81 | 29.20±5.70 |
|  | Median | 23.585 | 31.17 | 24.275 | 30.915 | 25.08 | 30.28 |
|  | Min~Max | 15.38~29.34 | 20.72~40.00 | 17.47~30.76 | 20.26~36.45 | 18.70~31.60 | 20.36~35.13 |
|  | P-value | 0.0573* |  | 0.0640* |  | 0.0613* |  |
| Day8 | N | 14 | 8 | 14 | 8 | 14 | 8 |
|  | Mean±Std | 29.69±6.26 | 35.42±6.21 | 29.88±5.45 | 33.46±5.98 | 30.27±4.97 | 32.99±4.83 |
|  | Median | 29.37 | 38.885 | 29.735 | 34.76 | 29.58 | 34.975 |
|  | Min~Max | 19.28~40.00 | 24.69~40.00 | 20.28~40.00 | 23.31~40.00 | 22.68~40.00 | 24.00~37.42 |
|  | P-value | 0.0626 ¥ |  | 0.1674* |  | 0.2264* |  |
| Day11 | N | 13 | 8 | 13 | 8 | 13 | 8 |
|  | Mean±Std | 32.40±6.29 | 34.62 ±4.69 | 32.38±5.08 | 33.65±4.53 | 32.30±4.18 | 32.87±4.33 |
|  | Median | 31.11 | 33.24 | 33.34 | 32.295 | 32.34 | 32.435 |
|  | Min~Max | 24.17~40.00 | 29.30~40.00 | 25.67~40.00 | 28.49~40.00 | 26.40~40.00 | 28.13~40.00 |
|  | P-value | 0.3193 ¥ |  | 0.5713* |  | 0.7685* |  |
| Day15 | N | 13 | 8 | 13 | 8 | 13 | 8 |
|  | Mean±Std | 37.97±3.28 | 35.42±4.04 | 37.71±2.75 | 35.16±4.24 | 37.65±2.66 | 34.57±4.69 |
|  | Median | 40.00 | 34.225 | 38.80 | 33.725 | 37.36 | 33.015 |
|  | Min~Max | 31.29~40.00 | 31.37~40.00 | 32.42~40.00 | 30.87~40.00 | 32.68~40.00 | 29.51~40.00 |
|  | P-value | 0.1486 ¥ |  | 0.1629 ¥ |  | 0.1414 ¥ |  |
|  |  | Clevudine<br>(N=16) | Placebo<br>(N=8) | Clevudine<br>(N=16) | Placebo<br>(N=8) | Clevudine<br>(N=16) | Placebo<br>(N=8) |
| Day22 | N | 12 | 8 | 12 | 8 | 12 | 8 |
|  | Mean±Std | 39.13±2.04 | 39.59±1.15 | 39.07±2.26 | 38.75±1.93 | 38.83±2.28 | 37.23±2.89 |
|  | Median | 40.00 | 40.00 | 40.00 | 40.00 | 40.00 | 37.23 |
|  | Min~Max | 34.69~40.00 | 36.76~40.00 | 32.99~40.00 | 35.69~40.00 | 33.53~40.00 | 31.76~40.00 |
|  | P-value | 0.7564 ¥ |  | 0.3888 ¥ |  | 0.1388 ¥ |  |
| Day29 | N | 13 | 8 | 13 | 8 | 13 | 8 |
|  | Mean±Std | 38.27±3.32 | 39.27±2.06 | 38.70±3.16 | 38.79±2.55 | 38.34±3.40 | 38.56±2.56 |
|  | Median | 40.00 | 40.00 | 40.00 | 40.00 | 40.00 | 40.00 |
|  | Min~Max | 31.43~40.00 | 34.16~40.00 | 31.50~40.00 | 32.91~40.00 | 30.77~40.00 | 33.42~40.00 |
|  | P-value | 0.4924 ¥ |  | 0.7917 ¥ |  | 0.6829 ¥ |  |
| Day4 - Baseline<br>(Change) | N | 16 | 8 | 16 | 8 | 16 | 8 |
|  | Mean±Std | 5.12±4.05 | 5.98±5.21 | 5.23±3.60 | 4.17±4.69 | 4.73±3.76 | 4.07±4.32 |
|  | Median | 5.84 | 5.99 | 5.84 | 4.08 | 5.265 | 3.31 |
|  | Min~Max | -2.17~11.48 | -1.58~13.11 | -1.72~11.66 | -2.90~12.89 | -1.88~10.88 | -2.18~11.46 |
|  | P-value (within) | 0.0001# | 0.014# | <.0001# | 0.0400# | 0.0001# | 0.0323# |
|  | P-value (between) | 0.6597* |  | 0.5444* |  | 0.7003* |  |
| Day8 - Baseline<br>(Change) | N | 14 | 8 | 14 | 8 | 14 | 8 |
|  | Mean±Std | 11.77±5.29 | 11.10±4.20 | 10.70±4.93 | 8.53±4.23 | 9.98±4.41 | 7.85±3.63 |
|  | Median | 12.145 | 12.435 | 10.79 | 8.88 | 9.74 | 7.72 |
|  | Min~Max | -1.45~19.46 | 5.08~16.86 | -1.63~18.95 | 4.00~15.74 | -0.59~17.68 | 3.97~12.09 |
| | P-value (within) | <.0001# | 0.0001# | <.0001# | 0.0007# | <.0001# | 0.0143\$ |
|  | P-value (between) | 0.7614* |  | 0.3100* |  | 0.2613* |  |
|  |  | Clevudine | Placebo | Clevudine | Placebo | Clevudine | Placebo |
|  |  | (N=16) | (N=8) | (N=16) | (N=8) | (N=16) | (N=8) |
| Day11 - Baseline | N | 13 | 8 | 13 | 8 | 13 | 8 |
| (Change) | Mean±Std | 14.45±7.30 | 10.29±9.32 | 13.06±5.77 | 8.71±8.03 | 11.85±5.32 | 7.73±8.22 |
|  | Median | 13.79 | 8.965 | 12.66 | 8.375 | 11.56 | 6.71 |
|  | Min~Max | 5.18~27.74 | -2.69~28.13 | 4.91~23.32 | -2.56~19.06 | 3.98~20.57 | -3.33~22.01 |
|  | P-value (within) | <.0001# | 0.0168# | <.0001# | 0.0181# | <.0001# | 0.0325# |
|  | P-value (between) | 0.2673* |  | 0.1647* |  | 0.1762* |  |
| Day15 - Baseline | N | 13 | 8 | 13 | 8 | 13 | 8 |
| (Change) | Mean±Std | 20.02±6.22 | 11.09±7.46 | 18.39±5.26 | 10.23±6.58 | 17.20±5.20 | 9.43±6.84 |
|  | Median | 21.18 | 10.055 | 19.40 | 9.67 | 18.83 | 7.51 |
|  | Min~Max | 11.22~28.16 | -1.59~20.69 | 11.66~25.60 | -1.03~18.38 | 9.82~24.63 | -1.95~20.15 |
|  | P-value (within) | <.0001# | 0.004# | <.0001# | 0.0032# | <.0001# | 0.0059# |
|  | P-value (between) | 0.0080* |  | 0.0054* |  | 0.0082* |  |
| Day22 - Baseline | N | 12 | 8 | 12 | 8 | 12 | 8 |
| (Change) | Mean±Std | 21.43±5.07 | 15.27±6.58 | 19.84±4.27 | 13.81±6.36 | 18.42±4.48 | 12.09±6.51 |
|  | Median | 21.275 | 16.925 | 19.45 | 16.00 | 17.965 | 12.955 |
|  | Min~Max | 14.65~28.16 | 5.08~24.89 | 13.71~25.60 | 3.79~22.13 | 12.08~24.63 | 0.30~23.12 |
|  | P-value (within) | <.0001# | 0.0003# | <.0001# | 0.0005# | <.0001# | 0.0012# |
|  | P-value (between) | 0.0295* |  | 0.0202* |  | 0.0187* |  |
| Day29 - Baseline | N | 13 | 8 | 13 | 8 | 13 | 8 |
| (Change) | Mean±Std | 20.31±5.81 | 14.94±6.09 | 19.38±4.68 | 13.85±4.93 | 17.90±4.71 | 13.42±4.63 |
|  | Median | 19.27 | 16.925 | 19.24 | 15.44 | 16.73 | 13.01 |
|  | Min~Max | 11.30~28.16 | 5.08~22.29 | 11.21~25.60 | 5.75~19.26 | 10.51~24.63 | 7.17~21.51 |
|  | P-value (within) | <.0001# | 0.0002# | <.0001# | 0.0001# | <.0001# | 0.0001# |
|  | P-value (between) | 0.0577* |  | 0.0185* |  | 0.0464* |  |

Note) \*: Two sample t-test, ¥ : Wilcoxon's rank sum test, #: Paired t-test, \$: Wilcoxon's signed rank test

For patients who underwent randomization during the first 5 days after the onset of symptoms and administered clevudine, the change in Ct value of RdRP gene from baseline to Day 29 was significantly greater when compared to placebo (Table 6 and Figure 5; RdRP gene; Day 29 p=0.0565).

**Figure 5.**
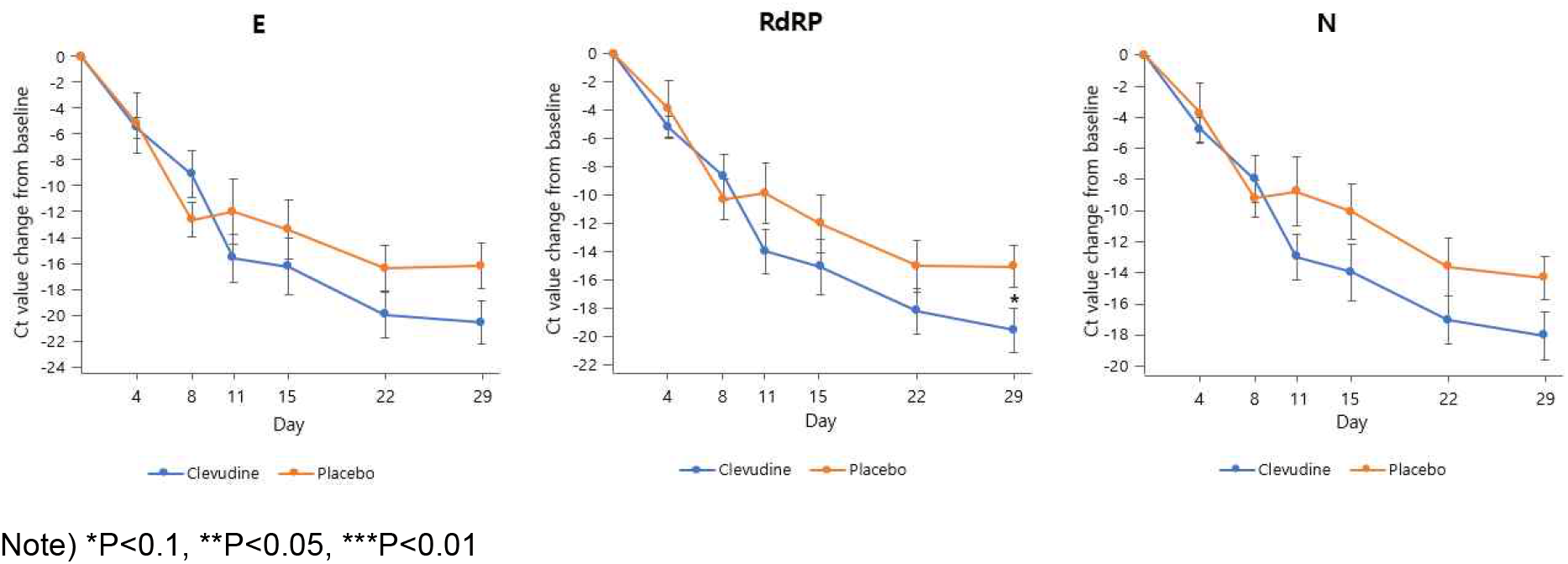
Change in Ct value from baseline (patients randomized within 5 days after symptom onset, ITT set)

**Table 6.**
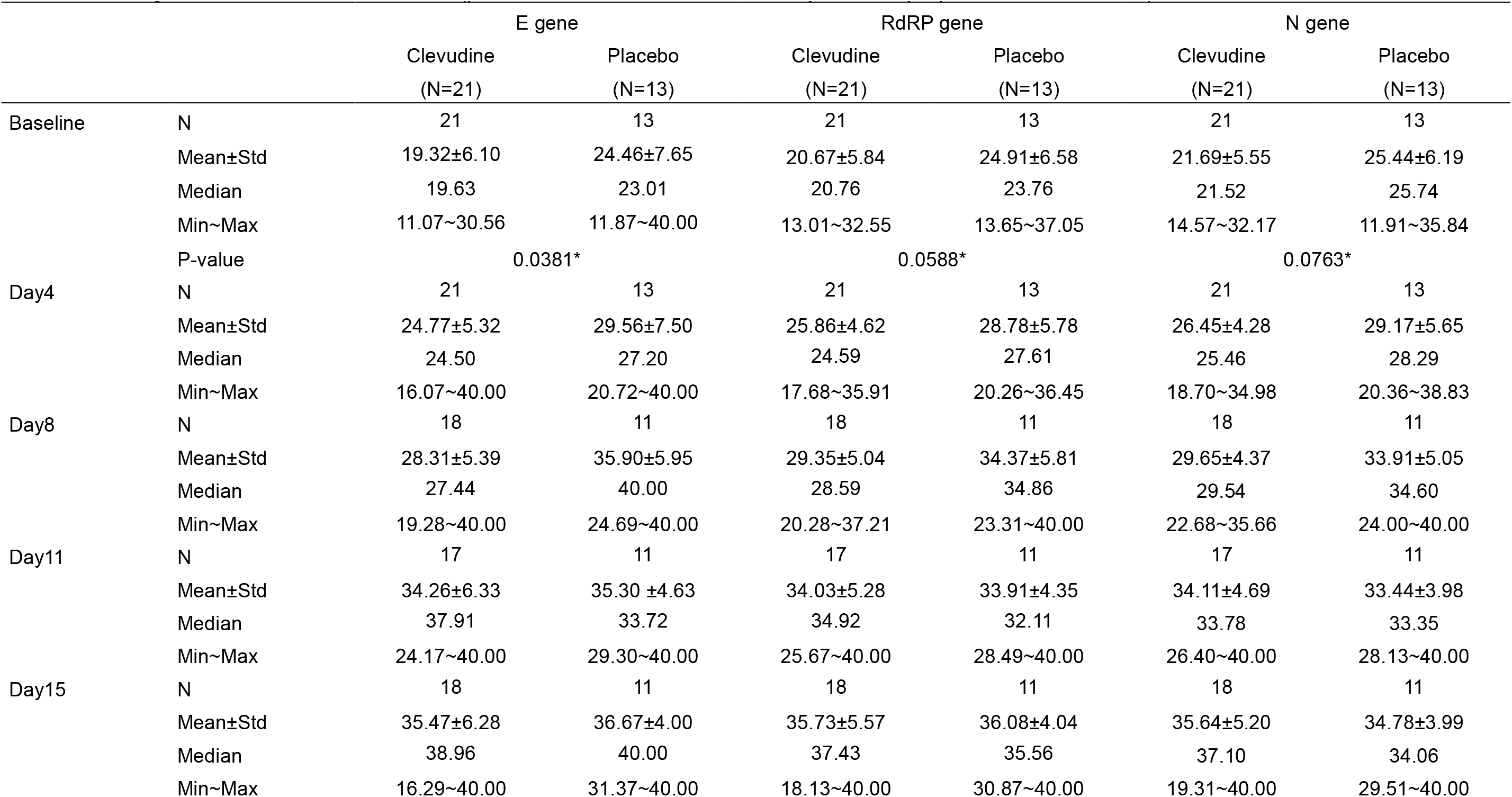
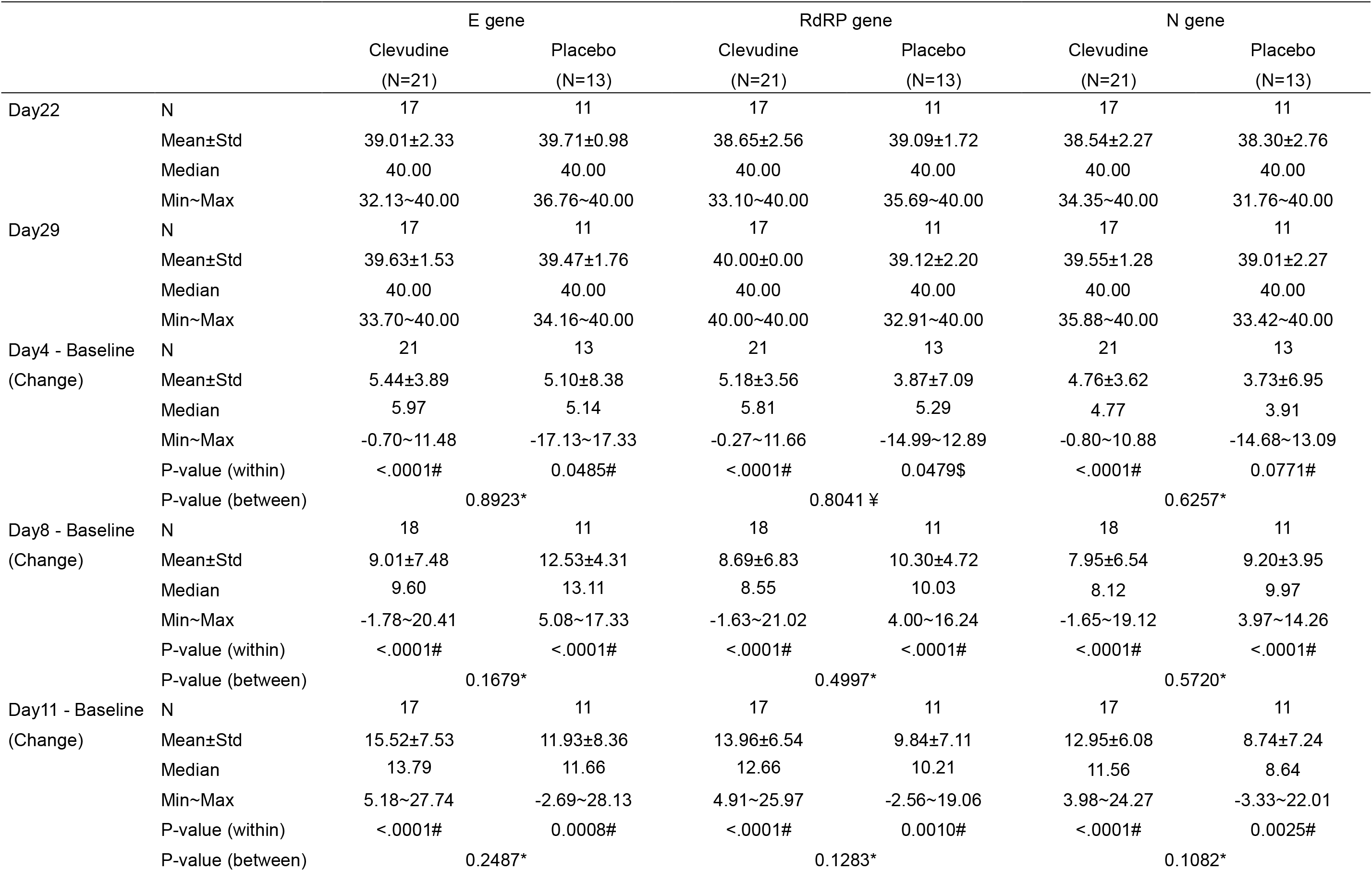
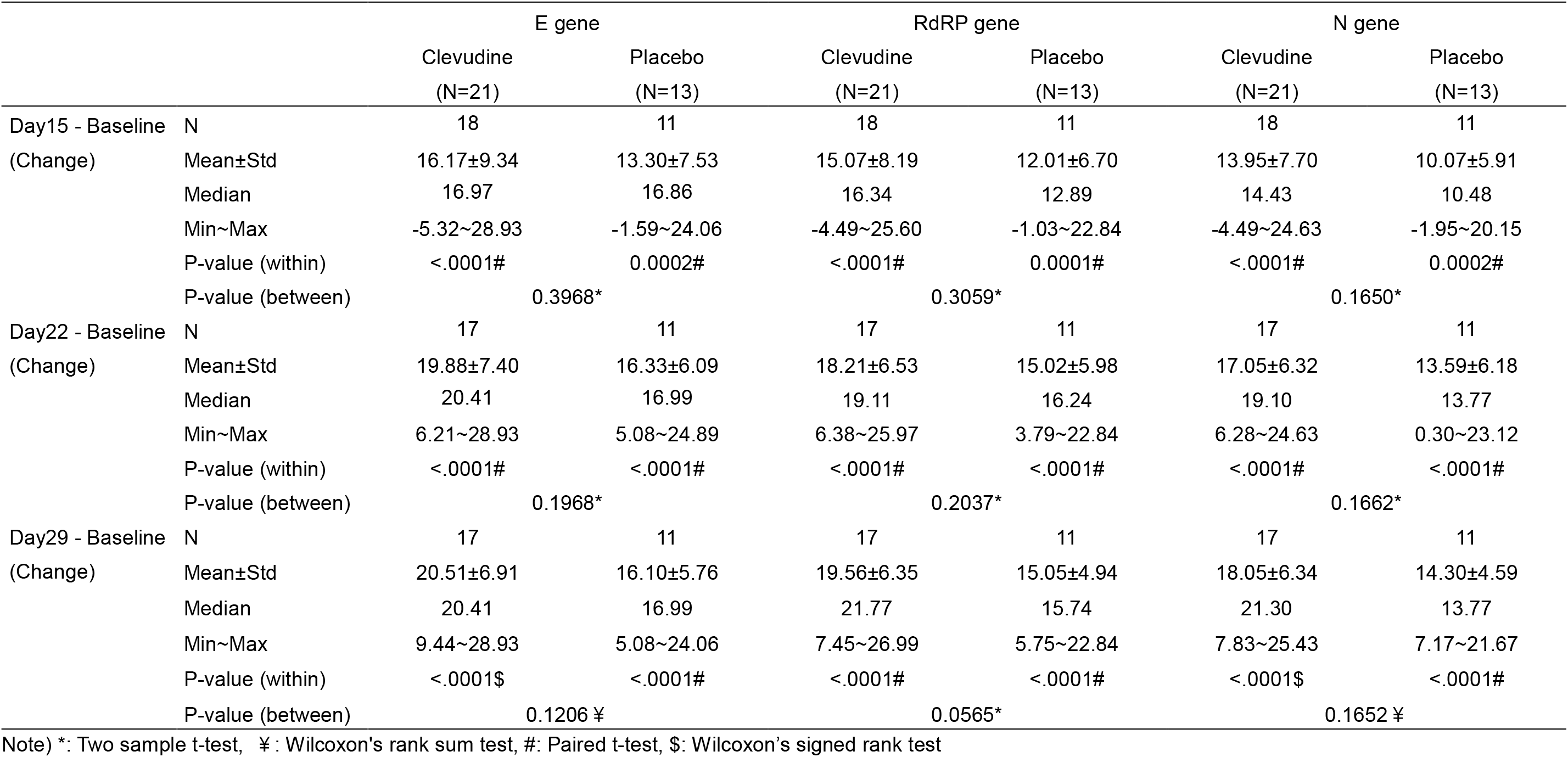
Change in Ct value from baseline (patients randomized within 5 days after symptom onset, ITT set)

In patients who underwent randomization during the first 7 days after the onset of symptoms and administered clevudine, the changes in Ct value of RdRP and N genes from baseline to Days 11 and 15 were significantly greater compared to placebo (Table 7 and Figure 6; RdRP gene; Day 11 p=0.0803, Day 15 p=0.0923; N gene; Day 11 p=0.0623, Day 15 p=0.0393).

**Figure 6.**
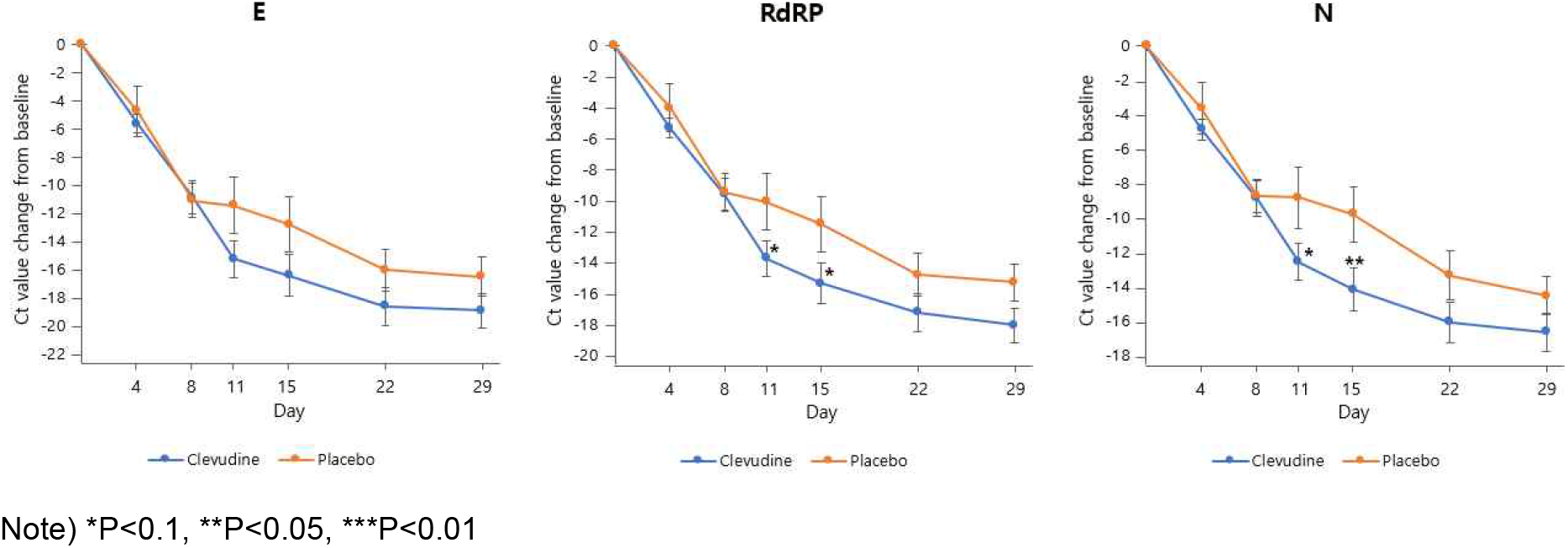
Change in Ct value from baseline (patients randomized within 7 days after symptom onset, ITT set)

**Table 7.** Change in Ct value from baseline (patients randomized within 7 days after symptom onset, ITT set)

|  |  | E gene |  | RdRP gene |  | N gene |  |
| --- | --- | --- | --- | --- | --- | --- | --- |
|  |  | Clevudine<br>(N=32) | Placebo<br>(N=17) | Clevudine<br>(N=32) | Placebo<br>(N=17) | Clevudine<br>(N=32) | Placebo<br>(N=17) |
| Baseline | N | 32 | 17 | 32 | 17 | 32 | 17 |
|  | Mean±Std | 20.18±5.67 | 24.57±7.06 | 21.25±5.47 | 25.14±5.99 | 22.30±5.31 | 25.69±5.66 |
|  | Median | 20.40 | 23.01 | 20.82 | 23.76 | 22.19 | 24.89 |
|  | Min~Max | 11.07~30.56 | 11.87~40.00 | 13.01~32.55 | 13.65~37.05 | 14.57~32.17 | 11.91~35.84 |
|  | P-value | 0.0221* |  | 0.0262* |  | 0.0427* |  |
| Day4 | N | 32 | 17 | 32 | 17 | 32 | 17 |
|  | Mean±Std | 25.79±5.81 | 29.28±7.14 | 26.55±4.86 | 29.14±5.89 | 27.15±4.50 | 29.31±5.35 |
|  | Median | 25.87 | 27.19 | 26.61 | 27.96 | 26.91 | 28.48 |
|  | Min~Max | 15.38~40.00 | 20.72~40.00 | 17.47~35.91 | 20.26~40.00 | 18.70~34.98 | 20.36~38.83 |
| Day8 | N | 29 | 15 | 29 | 15 | 29 | 15 |
|  | Mean±Std | 31.10±6.45 | 34.85±5.88 | 30.89±5.05 | 34.00±5.17 | 31.21±4.68 | 33.89±4.87 |
|  | Median | 31.30 | 37.77 | 32.88 | 34.72 | 32.58 | 34.60 |
|  | Min~Max | 19.28~40.00 | 24.69~40.00 | 20.28~40.00 | 23.31~40.00 | 22.68~40.00 | 24.00~40.00 |
| Day11 | N | 27 | 15 | 27 | 15 | 27 | 15 |
|  | Mean±Std | 35.25±5.91 | 35.21±4.37 | 34.80±4.96 | 34.61±4.31 | 34.75±4.46 | 34.01±3.92 |
|  | Median | 40.00 | 34.26 | 35.02 | 32.71 | 35.42 | 33.35 |
|  | Min~Max | 24.17~40.00 | 29.30~40.00 | 25.67~40.00 | 28.49~40.00 | 26.40~40.00 | 28.13~40.00 |
| Day15 | N | 28 | 15 | 28 | 15 | 28 | 15 |
|  | Mean±Std | 36.76±5.42 | 36.55±4.87 | 36.77±4.84 | 36.06±4.47 | 36.68±4.59 | 34.96±4.53 |
|  | Median | 40.00 | 40.00 | 39.40 | 37.09 | 37.46 | 35.11 |
|  | Min~Max | 16.29~40.00 | 24.90~40.00 | 18.13~40.00 | 26.91~40.00 | 19.31~40.00 | 26.24~40.00 |
|  |  | Clevudine<br>(N=32) | Placebo<br>(N=17) | Clevudine<br>(N=32) | Placebo<br>(N=17) | Clevudine<br>(N=32) | Placebo<br>(N=17) |
| Day22 | N | 26 | 15 | 26 | 15 | 26 | 15 |
|  | Mean±Std | 38.81±2.60 | 39.78±0.84 | 38.57±2.71 | 39.33±1.51 | 38.52±2.50 | 38.52±2.49 |
|  | Median | 40.00 | 40.00 | 40.00 | 40.00 | 40.00 | 40.00 |
|  | Min~Max | 31.40~40.00 | 36.76~40.00 | 32.82~40.00 | 35.69~40.00 | 32.75~40.00 | 31.76~40.00 |
| Day29 | N | 27 | 14 | 27 | 14 | 27 | 14 |
|  | Mean±Std | 39.17±2.42 | 39.58±1.56 | 39.38±2.25 | 39.31±1.97 | 39.05±2.53 | 39.18±2.02 |
|  | Median | 40.00 | 40.00 | 40.00 | 40.00 | 40.00 | 40.00 |
|  | Min~Max | 31.43~40.00 | 34.16~40.00 | 31.50~40.00 | 32.91~40.00 | 30.77~40.00 | 33.42~40.00 |
| Day4 - Baseline<br>(Change) | N | 32 | 17 | 32 | 17 | 32 | 17 |
|  | Mean±Std | 5.61±3.93 | 4.71±7.34 | 5.30±3.45 | 3.99±6.29 | 4.85±3.51 | 3.62±6.07 |
|  | Median | 6.35 | 4.73 | 5.84 | 5.01 | 4.66 | 4.06 |
|  | Min~Max | -2.17~12.16 | -17.13~17.33 | -1.72~11.66 | -14.99~12.89 | -1.88~10.92 | -14.68~13.09 |
| | P-value (within) | <.0001# | 0.0038\$ | <.0001# | 0.0079\$ | <.0001# | 0.0056\$ |
|  | P-value (between) | 0.7368 ¥ |  | 0.4948 ¥ |  | 0.4817 ¥ |  |
| Day8 - Baseline<br>(Change) | N | 29 | 15 | 29 | 15 | 29 | 15 |
|  | Mean±Std | 10.85±6.46 | 11.07±4.76 | 9.59±5.86 | 9.44±4.64 | 8.84±5.62 | 8.70±3.74 |
|  | Median | 12.16 | 12.54 | 9.41 | 9.66 | 8.61 | 9.48 |
|  | Min~Max | -1.78~20.41 | 3.08~17.33 | -1.63~21.02 | 3.27~16.24 | -1.65~19.12 | 3.86~14.26 |
|  | P-value (within) | <.0001# | <.0001# | <.0001# | <.0001# | <.0001# | <.0001# |
|  | P-value (between) | 0.9090* |  | 0.9326* |  | 0.9310* |  |
|  |  | Clevudine<br>(N=32) | Placebo<br>(N=17) | Clevudine<br>(N=32) | Placebo<br>(N=17) | Clevudine<br>(N=32) | Placebo<br>(N=17) |
| Day11 - Baseline | N | 27 | 15 | 27 | 15 | 27 | 15 |
| (Change) | Mean±Std | 15.21±6.79 | 11.43±7.81 | 13.70±5.94 | 10.05±6.94 | 12.55±5.54 | 8.82±6.85 |
|  | Median | 13.99 | 11.66 | 12.05 | 10.21 | 11.22 | 8.64 |
|  | Min~Max | 5.18~27.74 | -2.69~28.13 | 4.91~25.97 | -2.56~19.06 | 3.98~24.27 | -3.33~22.01 |
|  | P-value (within) | <.0001# | <.0001# | <.0001# | <.0001# | <.0001# | 0.0002# |
|  | P-value (between) | 0.1086* |  | 0.0803* |  | 0.0623* |  |
| Day15 - Baseline | N | 28 | 15 | 28 | 15 | 28 | 15 |
| (Change) | Mean±Std | 16.41±7.85 | 12.77±7.54 | 15.33±7.00 | 11.50±6.84 | 14.17±6.59 | 9.77±6.19 |
|  | Median | 15.56 | 16.86 | 15.01 | 12.89 | 13.32 | 10.48 |
|  | Min~Max | -5.32~28.93 | -1.59~24.06 | -4.49~25.60 | -1.03~22.84 | -4.49~24.63 | -1.95~20.15 |
|  | P-value (within) | <.0001# | <.0001# | <.0001# | <.0001# | <.0001# | <.0001# |
|  | P-value (between) | 0.1498* |  | 0.0923* |  | 0.0393* |  |
| Day22 - Baseline | N | 26 | 15 | 26 | 15 | 26 | 15 |
| (Change) | Mean±Std | 18.57±6.88 | 16.00±5.79 | 17.21±6.16 | 14.77±5.42 | 16.04±6.03 | 13.34±5.52 |
|  | Median | 18.83 | 16.99 | 17.12 | 16.24 | 15.69 | 13.77 |
|  | Min~Max | 6.21~28.93 | 5.08~24.89 | 6.38~25.97 | 3.79~22.84 | 5.53~24.63 | 0.30~23.12 |
|  | P-value (within) | <.0001# | <.0001# | <.0001# | <.0001# | <.0001# | <.0001# |
|  | P-value (between) | 0.2314* |  | 0.2114* |  | 0.1618* |  |
| Day29 - Baseline | N | 27 | 14 | 27 | 14 | 27 | 14 |
| (Change) | Mean±Std | 18.89±6.29 | 16.47±5.13 | 18.05±5.95 | 15.27±4.39 | 16.64±5.86 | 14.51±4.05 |
|  | Median | 17.90 | 17.16 | 17.38 | 15.99 | 15.33 | 14.69 |
|  | Min~Max | 9.44~28.93 | 5.08~24.06 | 7.45~26.99 | 5.75~22.84 | 7.83~25.43 | 7.17~21.67 |
| | P-value (within) | <.0001\$ | <.0001\$ | <.0001# | <.0001# | <.0001\$ | <.0001# |
|  | P-value (between) | 0.5362 ¥ |  | 0.1319* |  | 0.4747 ¥ |  |

Note) \*: Two sample t-test, ¥ : Wilcoxon's rank sum test, #: Paired t-test, \$: Wilcoxon's signed rank test

### Proportion of patients who showed improvement in lung involvement

A total of 43 patients had lung involvement at baseline, of which 28 and 15 patients were randomized in clevudine and placebo group, respectively. We evaluated the proportion of patients who showed improvement in lung invasion at Day 29. There was no significant difference in the proportion of patients who showed improvement in lung involvement between clevudine group (82.14%, 23/28) and placebo group (100%, 15/15) at Day 29 (Table 8).

**Table 8.** Proportion of patients who showed improvement in lung involvement (ITT set)

|  |  | Clevudine<br>(N=41) | Placebo<br>(N=20) | Total<br>(N=61) |
| --- | --- | --- | --- | --- |
| Day29(EOT) | Improvement | 23(82.14%) | 15(100.00%) | 38(88.37%) |
|  | No change or Aggravation | 5(17.86%) | 0(0.00%) | 5(11.63%) |
|  | Total | 28(65.12%) | 15(34.88%) | 43(100.00%) |
|  | P-value | 0.1449‡ | . | . |

Note) ‡: Fisher's exact test

### Proportion of patients with normal body temperature and normal oxygen saturation

The proportion of patients with normal body temperature and the proportion of patients with normal oxygen saturation at Day 4, 8, 11, 15, 22, and 29 were evaluated, but there was no significant difference between clevudine group and placebo group at every visit (Table 9, 10).

**Table 9.** Proportion of patients with normal body temperature (ITT set)

|  |  | Clevudine<br>(N=41) | Placebo<br>(N=20) | Total<br>(N=61) |
| --- | --- | --- | --- | --- |
| Day4 | Normal | 26(72.22%) | 14(82.35%) | 40(75.47%) |
|  | Abnormal | 10(27.78%) | 3(17.65%) | 13(24.53%) |
|  | Total | 36(67.92%) | 17(32.08%) | 53(100.00%) |
|  | P-value | 0.5112‡ | . | . |
| Day8 | Normal | 21(63.64%) | 11(73.33%) | 32(66.67%) |
|  | Abnormal | 12(36.36%) | 4(26.67%) | 16(33.33%) |
|  | Total | 33(68.75%) | 15(31.25%) | 48(100.00%) |
|  | P-value | 0.5089† | . | . |
| Day11 | Normal | 17(60.71%) | 12(80.00%) | 29(67.44%) |
|  | Abnormal | 11(39.29%) | 3(20.00%) | 14(32.56%) |
|  | Total | 28(65.12%) | 15(34.88%) | 43(100.00%) |
|  | P-value | 0.3084‡ | . | . |
| Day15 | Normal | 22(73.33%) | 11(73.33%) | 33(73.33%) |
|  | Abnormal | 8(26.67%) | 4(26.67%) | 12(26.67%) |
|  | Total | 30(66.67%) | 15(33.33%) | 45(100.00%) |
|  | P-value | >.9999‡ | . | . |
| Day22 | Normal | 23(95.83%) | 13(92.86%) | 36(94.74%) |
|  | Abnormal | 1(4.17%) | 1(7.14%) | 2(5.26%) |
|  | Total | 24(63.16%) | 14(36.84%) | 38(100.00%) |
|  | P-value | >.9999‡ | . | . |
| Day29 | Normal | 18(94.74%) | 10(100.00%) | 28(96.55%) |
|  | Abnormal | 1(5.26%) | 0(0.00%) | 1(3.45%) |
|  | Total | 19(65.52%) | 10(34.48%) | 29(100.00%) |
|  | P-value | >.9999‡ | . | . |

Note) †: Pearson's chi-square test, ‡: Fisher's exact test

**Table 10.**
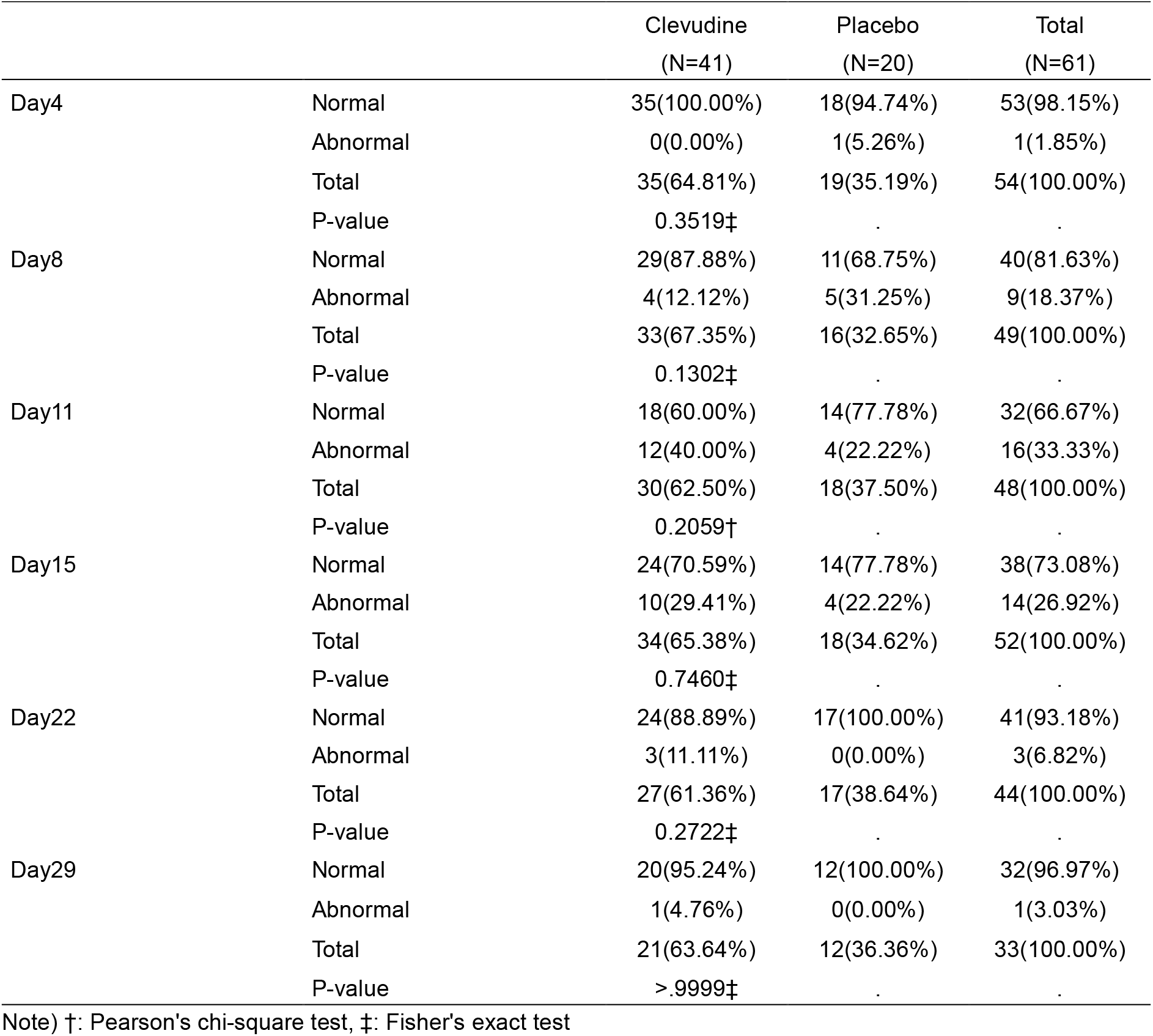
Proportion of patients with normal oxygen saturation (ITT set)

### Changes in CRP from baseline

Changes in CRP from baseline to Day 4, 8, 15, 22, and 29 were evaluated. Clevudine group showed greater decrease in CRP from baseline to all visits than placebo group, but there was no significant difference between clevudine group and placebo group at every visit (Table 11).

**Table 11.**
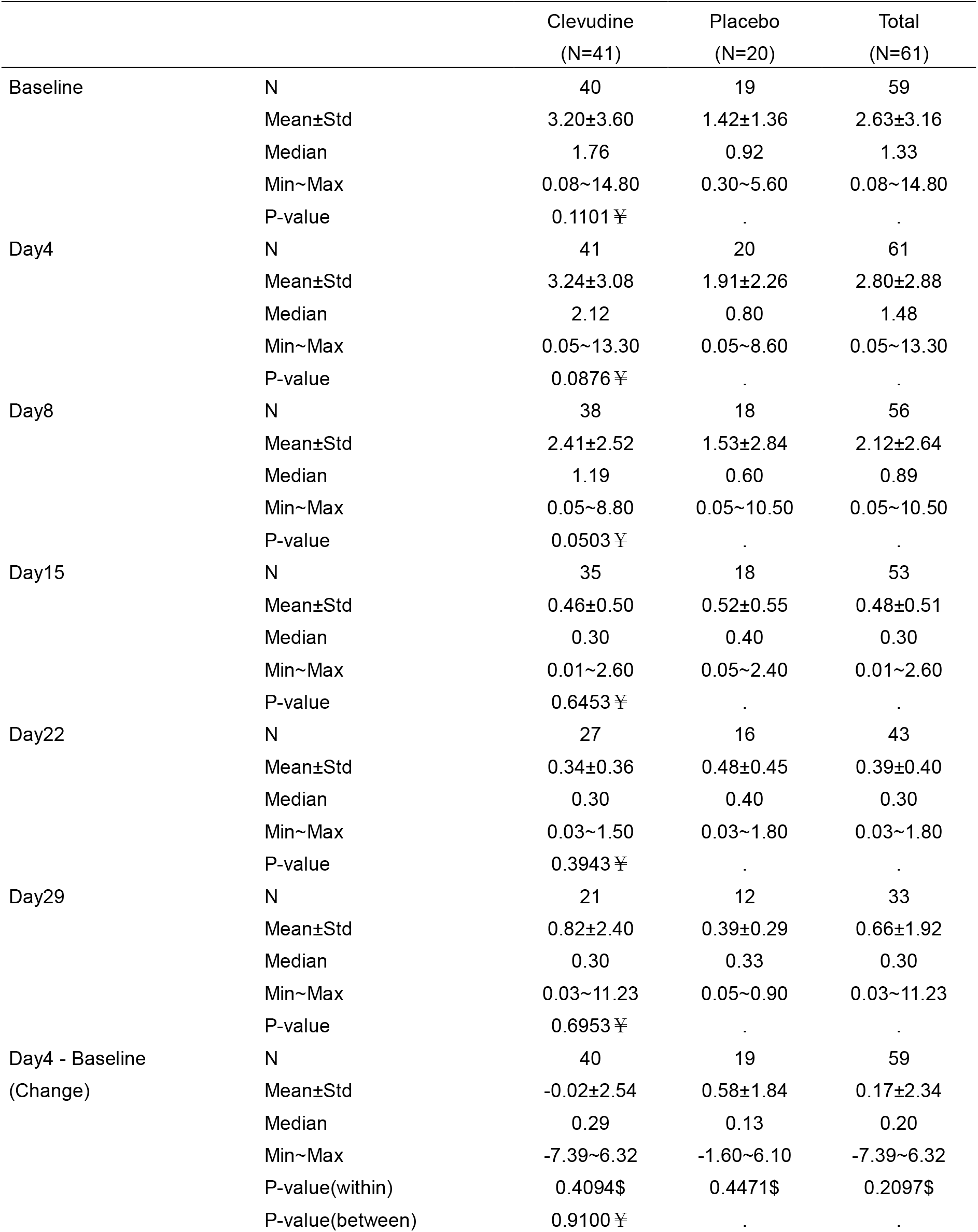
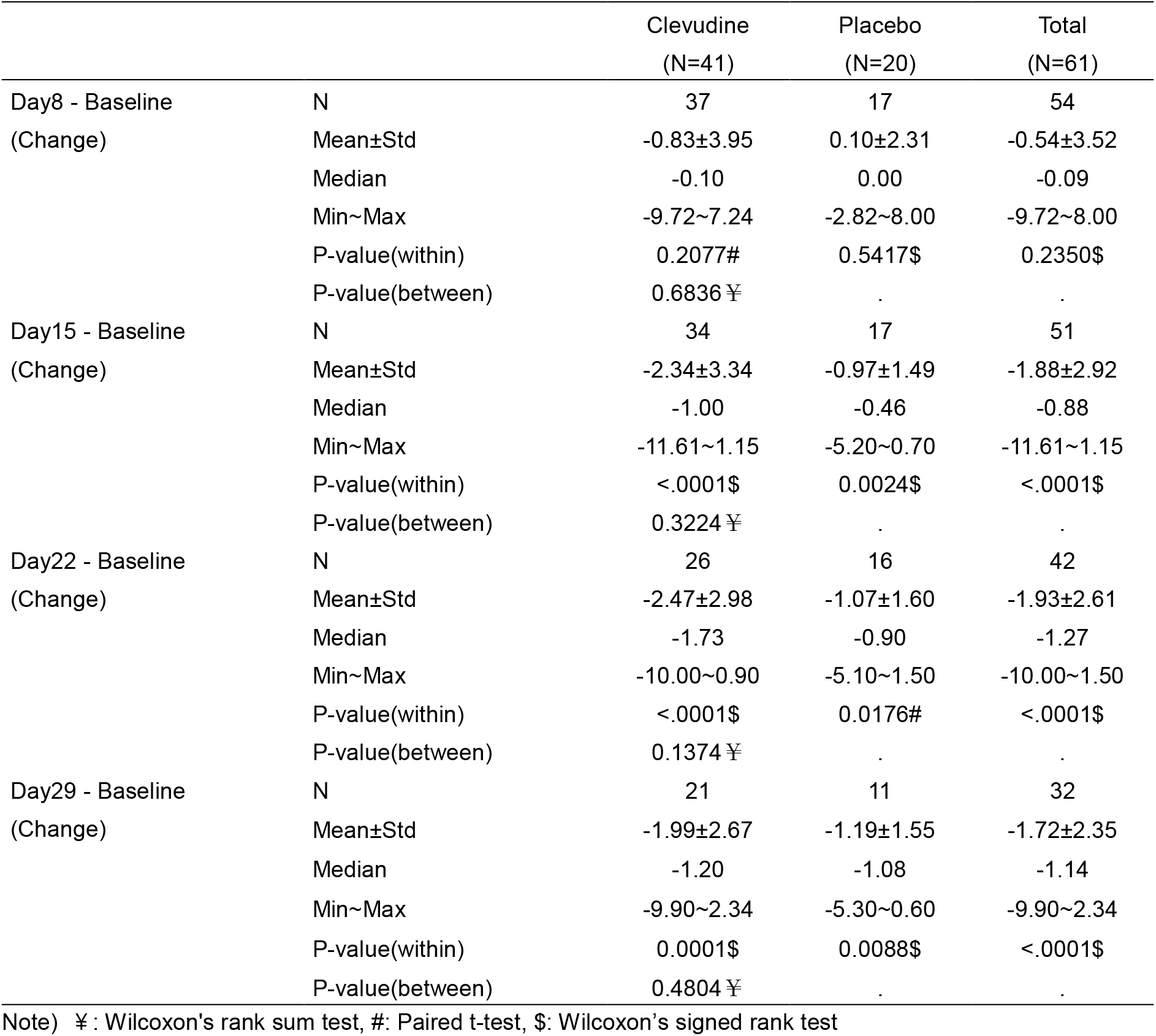
Change in CRP from baseline (ITT set)

### Safety and Tolerability

Clevudine and placebo were related with few and low-grade AEs. The incidence of ADRs were 34.15% (14/41) in clevudine group and 30.00% (6/20) in placebo group. There was no significant difference in ADRs occurrence between clevudine group and placebo group (p=0.7461). Five AEs led to discontinuation of clevudine and withdrawal from the study in five patients compared with one AE in one patient for placebo group. Grade 3 or higher AE occurred only in 1.64% (1/61, hypokalaemia) in the clevudine group and it was resolved. There was no serious AE or fatal outcome in this study. There were no marked differences in results of vital signs, physical examination, electrocardiogram, and laboratory tests between the treatment groups over the 29 days.

## Discussion

The current study found that clevudine was well tolerated but had no significant difference in the proportion of patients with negative result and the clinical efficacy outcomes compared to placebo in COVID-19 patients with moderate severity. However, clevudine was related with antiviral efficacy as evidenced by significant Ct value changes from baseline in patients with moderate COVID-19 and hypertension. The mechanism underlying this efficacy remains unclear. Furthermore, it is reported that antihypertensive drugs and glucose-lowering agents might positively influence the clinical outcomes and disease severity of COVID-19 and the degree of impact is different depending on the class of antihypertensive drugs and glucose-lowering agents according to observational retrospective studies.^32-34^ Considering these points, there is a need to draw a firm conclusion considering the effect of patient’s underlying diseases and concomitant medication on this result.

In patients randomized within the first 5 and 7 days after onset of symptoms, the greater change from baseline on Ct values observed in the clevudine group than the placebo group supports the finding that early treatment with clevudine would have an antiviral effect on SARS-CoV-2. Furthermore, more robust antiviral efficacy of clevudine can be expected in consideration of remarkably higher viral load in clevudine group than in placebo group at baseline.

With regard to safety profile, clevudine was well tolerated with no notable significant differences in ADRs or serious AEs between clevudine and placebo. There were no findings on clevudine that were new or not previously known from established safety information in HBV patients.

Based on the results from this study, the clinical use of clevudine in COVID-19 patients is expected to have a favorable outcome by suppressing the infectious virus replication, lowering viral load, and preventing the spread of COVID-19 in early stages of the infection.

## Conclusion

This is the first clinical study to compare the antiviral efficacy and safety of clevudine to placebo in Korean COVID-19 patients with moderate severity. The study has demonstrated a possible favorable outcome for the reduction of SARS-CoV-2 replication, with acceptable safety profile, when COVID-19 patients were treated with clevudine. This study further suggested that clevudine can be a favorable treatment option in the clinical practice for patients with moderate COVID-19, especially patients with hypertension. When we designed this study immediately after COVID-19 outbreak, the epidemiological and clinical features of COVID-19 had not been fully understood, therefore, we designed it as an initial therapeutic exploratory study. For this reason, this study had several limitations related to the study design (e.g., single blinded and small patient population, virological assessment as a primary outcome) and an imbalance in viral load at baseline between two treatment groups. Therefore, further large-scale clinical studies, preferably with various clinical endpoints and virus titer evaluation, are required to better understand the effectiveness of using clevudine in COVID-19 treatment. However, it is not simple to show a significant effectiveness of antiviral treatments in COVID-19 patients with mild or moderate severity because most of these patients do recover naturally from COVID-19 without any treatments.^35^ Considering this point, we need to design a clinical study aiming for reducing clinical risk of COVID-19 in mild to moderate patients at risk of severe illness when conducting further study.

## Supporting information

Disclosure of Interest

Research reporting checklist

## Data Availability

All data produced in the present study are available upon reasonable request to the authors.

